# Intravascular Imaging-Guided Versus Angiography-Guided PCI: Systematic Review and Meta-analysis of 24 Randomized Controlled Trials

**DOI:** 10.1101/2023.10.23.23297425

**Authors:** David Hong, Woochan Kwon, Seung Hun Lee, Doosup Shin, Hyun Kuk Kim, Kwan Yong Lee, Eun Ho Choo, Sung Eun Kim, Young Joon Hong, Young Keun Ahn, Myung Ho Jeong, Junho Ha, Minseok Hong, Ki Hong Choi, Taek Kyu Park, Jeong Hoon Yang, Young Bin Song, Joo-Yong Hahn, Seung-Hyuk Choi, Hyeon-Cheol Gwon, Joo Myung Lee, The RENOVATE COMPLEX-PCI Investigators

## Abstract

**Objective:** To evaluate comparative prognosis between intravascular imaging-guided PCI and angiography-guided PCI using a comprehensive meta-analysis including all previous randomized controlled trials.

**Design:** Systematic review and meta-analysis of randomized controlled trials.

**Data Sources:** Target trials were selected by a systematic electronic search strategy in the PubMed, EMBASE, and Cochrane databases from their inception to Sep, 2023.

**Study Selection:** Published randomized controlled trials which compared clinical outcomes between intravascular imaging-guided and angiography-guided PCI were included.

**Main Outcome and Measures:** Major adverse cardiac events (MACE), all-cause death, myocardial infarction (MI), target vessel revascularization (TVR), and stent thrombosis.

**Review Methods:** Random-effects model was used to calculate pooled relative risk (RR) and 95% confidence interval (CI) between intravascular imaging-guided and angiography-guided PCI. Heterogeneity was assessed by I^2^ values.

**Results:** Among a total of 14,037 patients (7,383 in intravascular imaging-guided PCI and 6,654 in angiography-guided PCI groups), intravascular imaging-guided PCI was associated with a lower risk of MACE than angiography-guided PCI (RR, 0.75; 95% CI, 0.67 to 0.84; P<0.001; I^2^, 18.9%), driven by lower risk of MI (RR, 0.83; 95% CI, 0.70 to 0.97; P=0.019; I^2^, 0.0%), TVR (RR, 0.70; 95% CI, 0.61 to 0.79; P<0.001; I^2^, 0.0%), and stent thrombosis (RR, 0.50; 95% CI, 0.32 to 0.78; P=0.002; I^2^, 0.0%). There was no difference in the risk of all-cause death (RR, 0.82; 95% CI, 0.67 to 1.01; P=0.068; I^2^, 0.0%), however, trials with 2^nd^ generation drug-eluting stent (DES) showed significant reduction of all-cause death following intravascular imaging-guided PCI (RR, 0.77; 95% CI, 0.61 to 0.95; P=0.018; I^2^, 0.0%).

**Conclusions:** Compared to angiography-guided PCI, intravascular imaging-guided PCI was associated with a reduced the risk of MACE, by lowering the risks of MI, TVR, and stent thrombosis. Pooled analysis of trials with 2^nd^ generation DES showed significantly lower risk of all-cause death following intravascular imaging-guided PCI than angiography-guided PCI.

**Systematic Review Registration:** PROSPERO, CRD42023402677

**Summary Boxes:** *What is already known on this topic:*

- Due to limited sample size, previous randomized controlled trials (RCTs) were not able to show significant differences in the risks of hard clinical events such as death, myocardial infarction (MI), or stent thrombosis between intravascular imaging-guided percutaneous coronary intervention (PCI) and angiography-guided PCI.
- Although previous meta-analyses showed that intravascular imaging-guided PCI resulted in a lower risk of death or MI than angiography-guided PCI, inclusion of both observational studies and RCTs caused heterogeneity and lowered the evidence level of the results.

*What this study adds:*

- The current meta-analysis exclusively included 24 RCTs and showed that intravascular imaging-guided PCI significantly reduced the risk of major adverse cardiac events (MACE), compared with angiography-guided PCI.
- It should be noted that intravascular imaging-guided PCI reduced not only the risk of target-vessel revascularization but also the risks of hard clinical events such as MI and stent thrombosis.

## Introduction

Percutaneous coronary intervention (PCI) is one of the important treatment options for patients with coronary artery stenosis. Despite improvement of drug-eluting stent (DES) platforms and PCI techniques, it is still unclear whether PCI can reduce death or myocardial infarction (MI) compared with guideline-directed medical treatment alone,^1^ ^2^ and PCI for complex coronary artery lesions is challenging in daily practice due to increased risk of stent failure.^3–5^ Therefore, continuous efforts have been made to find how to guide and optimize PCI to improve clinical outcomes. In this regard, intravascular imaging using intravascular ultrasound (IVUS) and optical coherence tomography (OCT) would be useful by providing information on lesion characteristics, reference vessel size, stent expansion and apposition, and acute complications.^6^

Previous randomized controlled trials (RCTs) have shown lower rates of major adverse cardiac events (MACE) after intravascular imaging-guided PCI than after angiography-guided PCI,^7–28^ although it mainly reduced the risk of repeat revascularization without significant difference in hard clinical events such as death, MI, or stent thrombosis. In contrast, the recently published RENOVATE-COMPLEX-PCI (Randomized Controlled Trial of Intravascular Imaging Guidance versus Angiography-Guidance on Clinical Outcomes After Complex Percutaneous Coronary Intervention) firstly demonstrated a superiority of intravascular imaging-guided PCI over angiography-guided PCI in patients with complex coronary artery lesions, mainly driven by reduced risk of cardiac death and MI with intravascular imaging guidance. The OCTOBER (European Trial on Optical Coherence Tomography Optimized Bifurcation Event Reduction) trial also showed the prognostic benefit of OCT-guided PCI over angiography-guided PCI in patients with complex bifurcation lesions.^29^ Conversely, ILUMIEN IV (OPtical Coherence Tomography [OCT] Guided Coronary Stent IMplantation Compared to Angiography: a Multicenter Randomized TriaL in PCI) failed to show significant difference in target vessel failure between the two strategies in patients with diabetes mellitus or complex coronary artery lesions.^30^

Therefore, we conducted an updated meta-analysis using all available RCTs to date to investigate whether intravascular imaging-guided PCI would improve clinical outcomes compared with angiography-guided PCI.

## Methods

### Data Sources and Searches

PubMed, EMBASE, Cochrane Central Register of Controlled Trials, the United States National Institutes of Health registry of clinical trials, and relevant websites were searched for pertinent published or unpublished studies from inception to September 2023. The electronic search strategy was complemented by manual examination of references cited by included articles, recent reviews, editorials, and meta-analyses. No restrictions were imposed on study period or sample size, but only articles in English were considered. Keywords included ‘intravascular ultrasound, ‘optical coherence tomography’, ‘PCI’, ‘percutaneous coronary intervention’, ‘coronary stenting’, ‘stenting’, ‘stent’, ‘stent implantation’, ‘angiography’, ‘impact’, ‘outcome’, ‘randomized’, and ‘randomized clinical trial’ (**Supplemental Table 1**). The present study complied with the Preferred Reporting Items for Systematic Reviews and Meta-Analyses (PRISMA) guidelines.^31^ The study protocol was prespecified and registered with PROSPERO (CRD42023402677). Institutional Review Board (IRB) of Samsung Medical Center waived IRB approval process based on the study design, which is the study-level meta-analysis using published materials.

### Study Selection

Studies were included if they met the following prespecified criteria: (1) RCTs comparing intravascular imaging-guided PCI versus angiography-guided PCI in patients with coronary artery disease; (2) clinical outcome assessment with follow-up duration of at least 1 month; and (3) clinical outcomes including all-cause death, cardiac death, MI, target vessel revascularization (TVR), or stent thrombosis. Non-randomized studies, duplicated reports of previous RCTs, or post-hoc analysis including cost-effectiveness analysis of reported RCTs were not included (**Supplemental Table 2**). Two independent investigators (D.H. and W.K.) screened titles and abstracts, identified duplicates, performed full-article reviews, and determined inclusion. Two other investigators (S.H.L. and J.M.L.) supervised the search and adjudicated disagreements.

### Data Extraction and Quality Assessment

Summary data as reported in the published manuscripts were analyzed. A standardized form was used to extract study characteristics; trial design; number of trial population; median follow-up duration; type of intravascular imaging devices; demographics; clinical and angiographic eligibility criteria, including clinical diagnosis and angiographic lesion characteristics; proportion of cardiovascular risk factors; and used stent types. The rates of all-cause mortality, MI, TVR, stent thrombosis, and MACE were collected, along with the definition of clinical events, as reported on an intention-to-treat basis. For trials which used both IVUS and OCT, group size and number of events were separately extracted, according to the type of intravascular imaging devices. In trials with available reports for both short- and long-term clinical outcomes, those with the longest follow-up duration were used in the current meta-analysis. In addition, unadjusted results of clinical events were used in the current meta-analysis.

The quality of eligible trials was assessed using the Cochrane Collaborations tool Version 2 for assessing the risk of bias in RCTs regarding random sequence generation, allocation concealment, blinding of participants and outcome assessment, incomplete outcome assessment, or selective reporting. We did not exclude individual studies from the analysis based on the risk of bias in RCTs.

### Outcomes and Definitions

The primary outcome was MACE at the longest available follow-up. Since the definition of MACE or composite outcomes varied across the trials, we used representative composite outcome reported under intention-to-treat principle in the included trials. The definition of MACE used in the included trials are presented in **Supplemental Table 3**. In most trials, MACE was a composite of all-cause death, MI, and TVR. Secondary outcomes included all-cause death, MI, TVR, and stent thrombosis.

### Data Synthesis and Analysis

Primary and secondary outcomes were analyzed using random-effects models. Relative risks (RRs) with 95% confidence intervals (CIs) were presented as summary statistics. Because all included studies showed heterogeneity regarding study protocol and populations, fixed-effects models were only used for sensitivity analyses to check whether these models yielded similar results. The pooled RRs and 95% confidence intervals were calculated using the DerSimonian and Laird method for random effects, as well as the Mantel–Haenszel method for fixed effects. Because of progressive changes in primary study designs and clinical practice patterns, especially type of used stents (bare metal stent, 1^st^ or 2^nd^ generation DES), we evaluated the impact of publication date on the overall effect of pooled RRs for the risk of MACE by a cumulative meta-analysis. Since all of the included trials in the cumulative meta-analysis had the same comparison groups, cumulative pooled effect estimates up to time point of last study inclusion could reflect temporal trends in effect size (RR). Statistical heterogeneity was quantified with the I^2^ statistics. I² values <25%, 25% to 50%, and >50% indicate low, moderate, and high degrees of heterogeneity, respectively. Small study bias (publication bias) was assessed by funnel plot asymmetry and Egger’s and Begg’s tests; when visual asymmetry of funnel plot was suspected, the trim-and-fill method was used to estimate the number of missing studies and to calculate a corrected RR, as if these studies were present. The influence of an individual study was explored by estimating pooled RRs, with stepwise exclusion of one study.

Subgroup analyses were performed to determine whether effects differed across subgroups. These subgroups analyses were analyzed: (1) type of intravascular imaging devices (IVUS or OCT); (2) angiographic characteristics (all-comers population or patients with complex coronary artery disease); (3) type of complex coronary artery lesions (chronic total occlusion, left main disease, or diffuse long lesion); (4) type of used stents (bare metal stent, 1^st^ or 2^nd^ generation DES); (5) clinical presentation (stable ischemic heart disease or acute coronary syndrome); and (6) presence of cardiovascular risk factors (diabetes mellitus or chronic kidney disease). Random-effects meta regression analysis was performed to evaluate differential effect size of intravascular imaging-guided PCI for MACE according to each subgroup. Two-sided P-values <0.05 were considered statistically significant. Statistical analysis was performed using STATA/SE 12.0 (Stata Corp LP, College Station, Texas, USA) and R programming language, version 4.2.2 (R Foundation for Statistical Computing, Vienna, Austria).

## Results

### Included Trials and Patient Characteristics

Of a total of 809 searched articles, 38 were retrieved for full article review and 24 met prespecified inclusion criteria and were finally included (**Figure 1**). A total of 14,037 patients from 24 RCTs were included in the current meta-analysis. Among the total patients, 7,383 (52.6%) patients were treated with intravascular imaging-guided PCI and the remaining 6,654 (47.4%) patients were with angiography-guided PCI. The characteristics of the included trials are summarized in **Table 1**. The study population of 11 trials were all-comers without restriction by angiographic lesion characteristics and the remaining 13 trials exclusively enrolled patients with complex coronary artery lesions. Fifteen trials exclusively used IVUS, 6 trials exclusively used OCT, and the other 3 trials used both IVUS and OCT. 17 trials used 2^nd^ generation DES and the other 7 trials used bare metal stent or 1^st^ generation DES. The follow-up duration of each trial ranged from 6 months to 5 years and weighted median follow-up duration in the current meta-analysis was 2.0 years.

**Figure 1.**
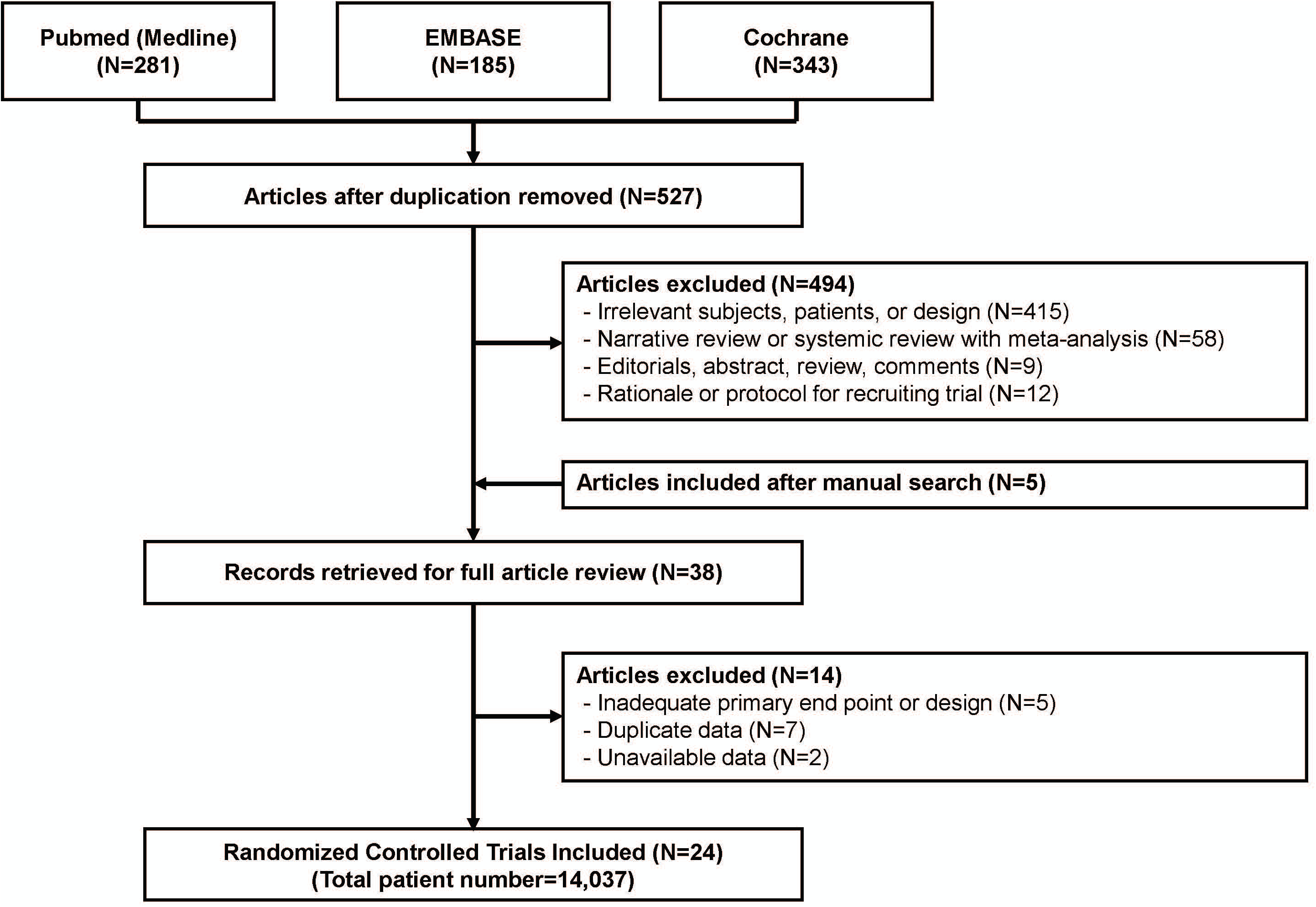
Flow Diagram of Study Selection. The study flow diagram was depicted following the guideline of Preferred Reporting Items for Systematic Reviews and Meta-Analyses (PRISMA).

**Table 1.**
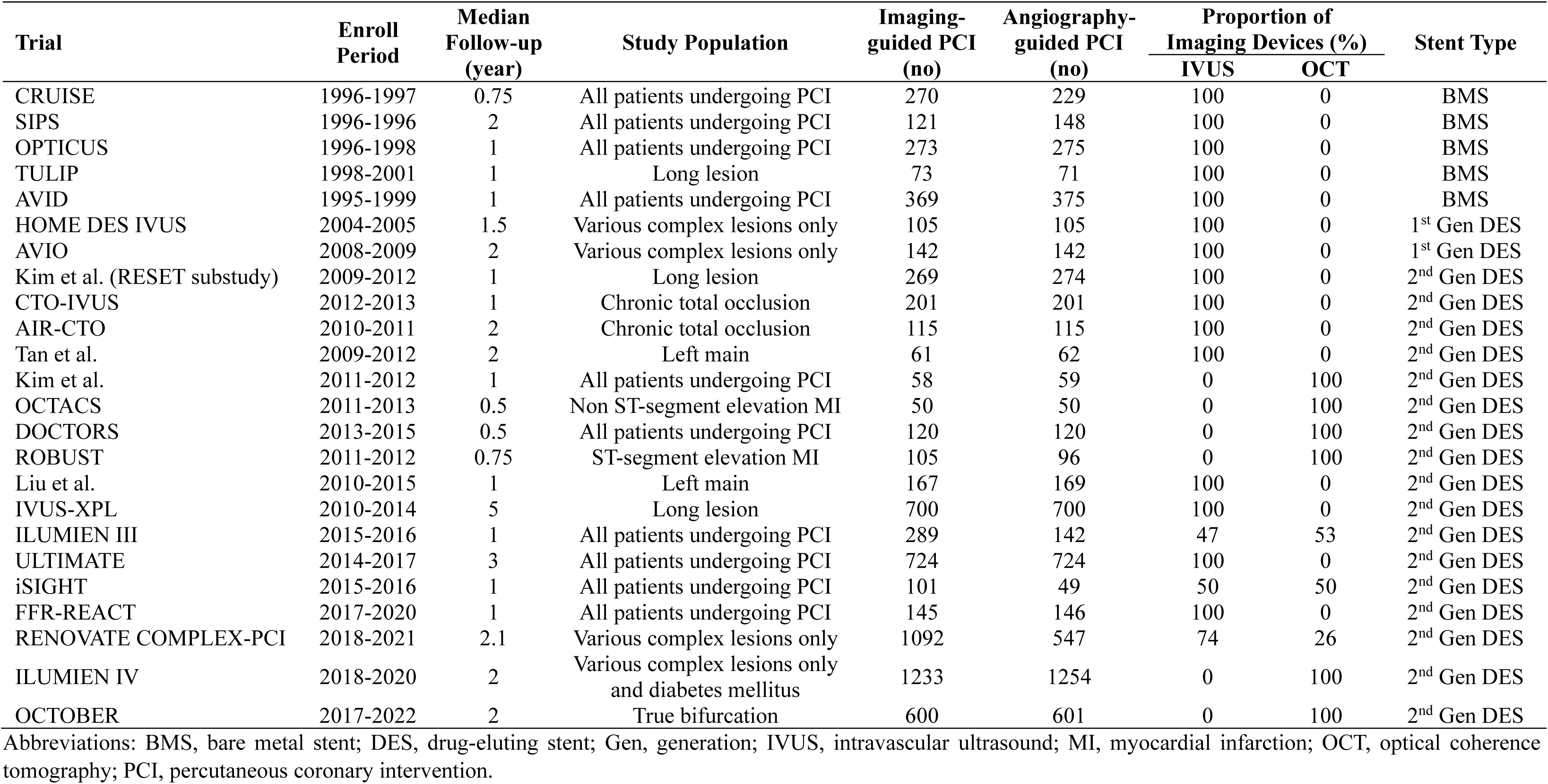
Baseline Characteristics of the Included Trials.

| Trial | Enroll Period | Median Follow-up (year) | Study Population | Imaging-guided PCI (no) | Angiography-guided PCI (no) | Proportion of Imaging Devices (%) |  | Stent Type |
| --- | --- | --- | --- | --- | --- | --- | --- | --- |
|  |  |  |  |  |  | IVUS | OCT |  |
| CRUISE | 1996-1997 | 0.75 | All patients undergoing PCI | 270 | 229 | 100 | 0 | BMS |
| SIPS | 1996-1996 | 2 | All patients undergoing PCI | 121 | 148 | 100 | 0 | BMS |
| OPTICUS | 1996-1998 | 1 | All patients undergoing PCI | 273 | 275 | 100 | 0 | BMS |
| TULIP | 1998-2001 | 1 | Long lesion | 73 | 71 | 100 | 0 | BMS |
| AVID | 1995-1999 | 1 | All patients undergoing PCI | 369 | 375 | 100 | 0 | BMS |
| HOME DES IVUS | 2004-2005 | 1.5 | Various complex lesions only | 105 | 105 | 100 | 0 | 1<sup>st</sup> Gen DES |
| AVIO | 2008-2009 | 2 | Various complex lesions only | 142 | 142 | 100 | 0 | 1<sup>st</sup> Gen DES |
| Kim et al. (RESET substudy) | 2009-2012 | 1 | Long lesion | 269 | 274 | 100 | 0 | 2<sup>nd</sup> Gen DES |
| CTO-IVUS | 2012-2013 | 1 | Chronic total occlusion | 201 | 201 | 100 | 0 | 2<sup>nd</sup> Gen DES |
| AIR-CTO | 2010-2011 | 2 | Chronic total occlusion | 115 | 115 | 100 | 0 | 2<sup>nd</sup> Gen DES |
| Tan et al. | 2009-2012 | 2 | Left main | 61 | 62 | 100 | 0 | 2<sup>nd</sup> Gen DES |
| Kim et al. | 2011-2012 | 1 | All patients undergoing PCI | 58 | 59 | 0 | 100 | 2<sup>nd</sup> Gen DES |
| OCTACS | 2011-2013 | 0.5 | Non ST-segment elevation MI | 50 | 50 | 0 | 100 | 2<sup>nd</sup> Gen DES |
| DOCTORS | 2013-2015 | 0.5 | All patients undergoing PCI | 120 | 120 | 0 | 100 | 2<sup>nd</sup> Gen DES |
| ROBUST | 2011-2012 | 0.75 | ST-segment elevation MI | 105 | 96 | 0 | 100 | 2<sup>nd</sup> Gen DES |
| Liu et al. | 2010-2015 | 1 | Left main | 167 | 169 | 100 | 0 | 2<sup>nd</sup> Gen DES |
| IVUS-XPL | 2010-2014 | 5 | Long lesion | 700 | 700 | 100 | 0 | 2<sup>nd</sup> Gen DES |
| ILUMIEN III | 2015-2016 | 1 | All patients undergoing PCI | 289 | 142 | 47 | 53 | 2<sup>nd</sup> Gen DES |
| ULTIMATE | 2014-2017 | 3 | All patients undergoing PCI | 724 | 724 | 100 | 0 | 2<sup>nd</sup> Gen DES |
| iSIGHT | 2015-2016 | 1 | All patients undergoing PCI | 101 | 49 | 50 | 50 | 2<sup>nd</sup> Gen DES |
| FFR-REACT | 2017-2020 | 1 | All patients undergoing PCI | 145 | 146 | 100 | 0 | 2<sup>nd</sup> Gen DES |
| RENOVATE COMPLEX-PCI | 2018-2021 | 2.1 | Various complex lesions only | 1092 | 547 | 74 | 26 | 2<sup>nd</sup> Gen DES |
| ILUMIEN IV | 2018-2020 | 2 | Various complex lesions only and diabetes mellitus | 1233 | 1254 | 0 | 100 | 2<sup>nd</sup> Gen DES |
| OCTOBER | 2017-2022 | 2 | True bifurcation | 600 | 601 | 0 | 100 | 2<sup>nd</sup> Gen DES |

Abbreviations: BMS, bare metal stent; DES, drug-eluting stent; Gen, generation; IVUS, intravascular ultrasound; MI, myocardial infarction; OCT, optical coherence tomography; PCI, percutaneous coronary intervention.

Regarding the quality of included trials, most trials were deemed to be at low risk of bias, although some criteria were unclearly described in few trials (**Supplemental Figure 1** and **Supplemental Table 4**). The characteristics of patients in the included trials are presented in **Supplemental Table 5**. Briefly, weighted mean age of the study population was 64.3 years and 75.6% were male. 64.8%, 30.5%, and 60.4% of the study population had hypertension, diabetes mellitus, and dyslipidemia, respectively. 55.7% of the study population presented with acute coronary syndrome. As only RCTs were included in the current study, baseline characteristics were balanced between intravascular imaging-guided PCI and angiography-guided PCI groups.

### Clinical Outcomes

Among 14,037 patients from 24 RCTs, observed rates of MACE in pooled analysis were 9.1% and 12.4% at a weighted median follow-up of 2.0 years for intravascular imaging-guided PCI and angiography-guided PCI groups, respectively. In random-effects model, intravascular imaging-guided PCI was associated with a lower risk of MACE than angiography-guided PCI (RR, 0.75; 95% CI, 0.67 to 0.84; P<0.001; heterogeneity I^2^, 18.9%) (**Figure 2**). Number needed to treat to prevent one MACE was 33 (95% CI, 25 to 50). A fixed-effects model yielded similar results (RR, 0.75; 95% CI, 0.68 to 0.83; P<0.001; heterogeneity I^2^, 18.9%). The lower risk of MACE in intravascular imaging-guided PCI was driven by significantly lower risks of MI (RR, 0.83; 95% CI, 0.70 to 0.97; P=0.019; heterogeneity I^2^, 0.0%) and TVR (RR, 0.70; 95% CI, 0.61 to 0.79; P<0.001; heterogeneity I^2^, 0.0%). Regarding stent thrombosis, observed rates were 0.5% and 1.2% at a weighted median follow-up of 2.0 years for intravascular imaging-guided PCI and angiography-guided PCI groups, respectively (RR, 0.50; 95% CI, 0.32 to 0.78; P=0.002; heterogeneity I^2^, 0.0%). However, there was no difference in the risk of all-cause death between the 2 groups (RR, 0.82; 95% CI, 0.67 to 1.01; P=0.068; heterogeneity I^2^, 0.0%) (**Figure 3**).

**Figure 2.**
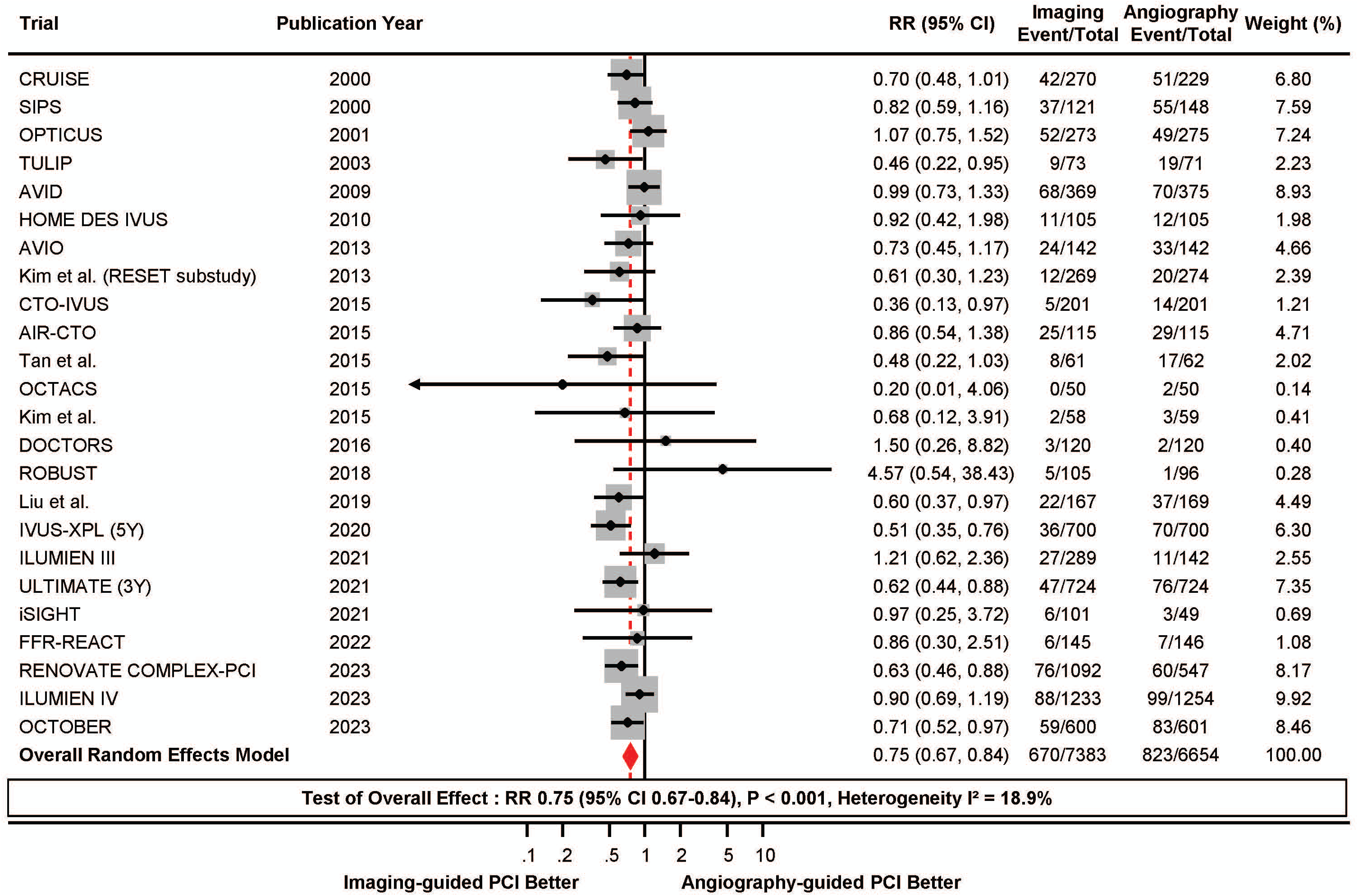
Meta-Analysis Comparing Major Adverse Cardiac Event Between Intravascular Imaging vs. Angiography-Guided PCI. Forest plots comparing major adverse cardiac event, a composite of all-cause death, myocardial infarction, and target vessel revascularization between intravascular imaging-guided and angiography-guided PCI. RRs with 95% CIs are displayed for individual studies and the pooled overall effect. Abbreviations: **AIR-CTO**, Angiographic and clinical comparisons of intravascular ultrasound-versus angiography-guided drug-eluting stent implantation for patients with chronic total occlusion lesions; **AVID**, Angiography Versus Intravascular ultrasound-Directed stent placement; **AVIO**, Angiography Vs. IVUS Optimization; **CRUISE**, Can Routine Ultrasound Influence Stent Expansion; **CI**, indicates confidence interval; **CTO-IVUS**, Chronic Total Occlusion InterVention with drUg-eluting Stents guided by IVUS; **DOCTORS**, Does Optical Coherence Tomography Optimize Results of Stenting; **FFR-REACT**, Fractional flow reserve guided percutaneous coronary intervention optimization directed by high-definition intravascular ultrasound versus standard of care; **HOME DES IVUS**, Long-Term Health Outcome and Mortality Evaluation After Invasive Coronary Treatment Using Drug Eluting Stents with or without the IVUS Guidance; **iSIGHT**, Optical Coherence Tomography Versus Intravascular Ultrasound and Angiography to Guide Percutaneous Coronary Interventions; **IVUS-XPL**, Impact of Intravascular Ultrasound Guidance on Outcomes of Xience Prime Stents in Long Lesions; **OCTACS**, Optical Coherence Tomography Guided Percutaneous Coronary Intervention With Nobori Stent Implantation in Patients With Non ST Segment Elevation Myocardial Infarction; **OCTOBER**, European Trial on Optical Coherence Tomography Optimized Bifurcation Event Reduction; **OPTICUS**, OPTimization with ICUS to reduce stent restenosis; **RENOVATE COMPLEX-PCI**, Randomized Controlled Trial of Intravascular Imaging Guidance versus Angiography-Guidance on Clinical Outcomes After Complex Percutaneous Coronary Intervention; **PCI**, percutaneous coronary intervention; **RESET**, Real Safety and Efficacy of a 3-Month Dual Antiplatelet Therapy Following Zotarolimus-Eluting Stents Implantation; **ROBUST**, Comparison of Biolimus A9 and Everolimus Drug-Eluting Stents in Patients With ST Segment Elevation Myocardial Infarction; **RR**, relative risk; **SIPS**, Strategy for Intracoronary Ultrasound-Guided PTCA and Stenting; **TULIP**, Thrombocyte activity evaluation and effects of Ultrasound guidance in Long Intracoronary stent Placement; and **ULTIMATE**, Intravascular Ultrasound Guided Drug Eluting Stents Implantation in “All-Comers” Coronary Lesions.

**Figure 3.**
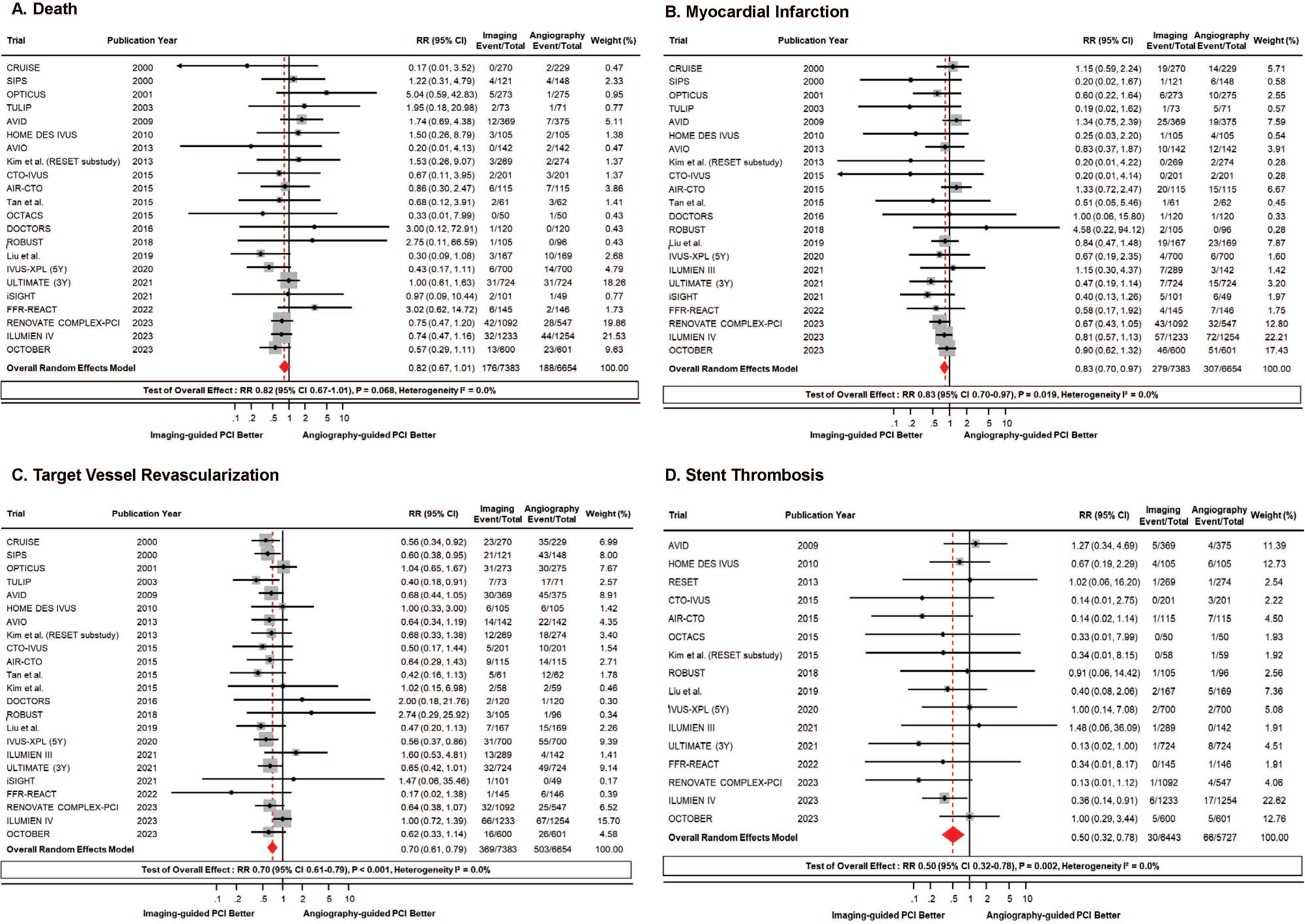
Meta-Analysis Comparing Clinical Outcomes Between Intravascular Imaging vs. Angiography-Guided PCI. Forest plots comparing individual clinical outcomes between intravascular imaging-guided and angiography-guided PCI. RRs with 95% CIs are displayed for individual studies and the pooled overall effect. (A) all-cause death, (B) myocardial infarction, (C) target vessel revascularization, and (D) stent thrombosis. Abbreviations are as in Figure 2.

To assess the robustness and bias of the main results, additional analyses were performed. Fixed-effects models yielded consistent results with those from the random-effects models for MI, TVR, and stent thrombosis. In the cumulative meta-analysis to evaluate the temporal trend, intravascular imaging-guided PCI consistently showed a lower risk of MACE than angiography-guided PCI over time from 2000 to 2023 (**Supplemental Figure 2**). No individual study substantially influenced the pooled effect estimate for MACE (**Supplemental Figure 3**). Funnel plots, supported by Egger’s and Begg’s tests, indicated no small study bias for MACE. The adjusted RRs for MACE by trim-and-fill method showed consistent results (RR, 0.75; 95% CI, 0.67 to 0.84; P<0.001) (**Supplemental Figure 4**).

### Subgroup Analysis

In subgroup analysis according to the all-comers population or patients with complex coronary artery lesions, intravascular imaging-guided PCI consistently showed a lower risk of MACE than angiography-guided PCI. However, the prognostic benefit of the intravascular imaging-guided PCI was relatively higher in patients treated by 2^nd^ generation DES than previous types of stents (Interaction P value=0.014) (**Figure 4**). The prognostic benefit of intravascular imaging-guided PCI over angiography-guided PCI was consistently observed in both stable ischemic heart disease and acute coronary syndrome (**Supplemental Figure 5**). Other subgroup analyses also showed similar results (**Figure 4**). Although intravascular imaging-guided PCI marginally reduced the risk of all-cause death in overall meta-analysis, subgroup of the trials with 2^nd^ generation DES showed significant reduction of all-cause death in intravascular imaging-guided PCI group than angiography-guided PCI group (RR, 0.77; 95% CI, 0.61 to 0.95; P=0.018; heterogeneity I^2^, 0.0%), which was not seen in the trials with bare metal stent or 1^st^ generation DES (**Figure 5**).

**Figure 4.**
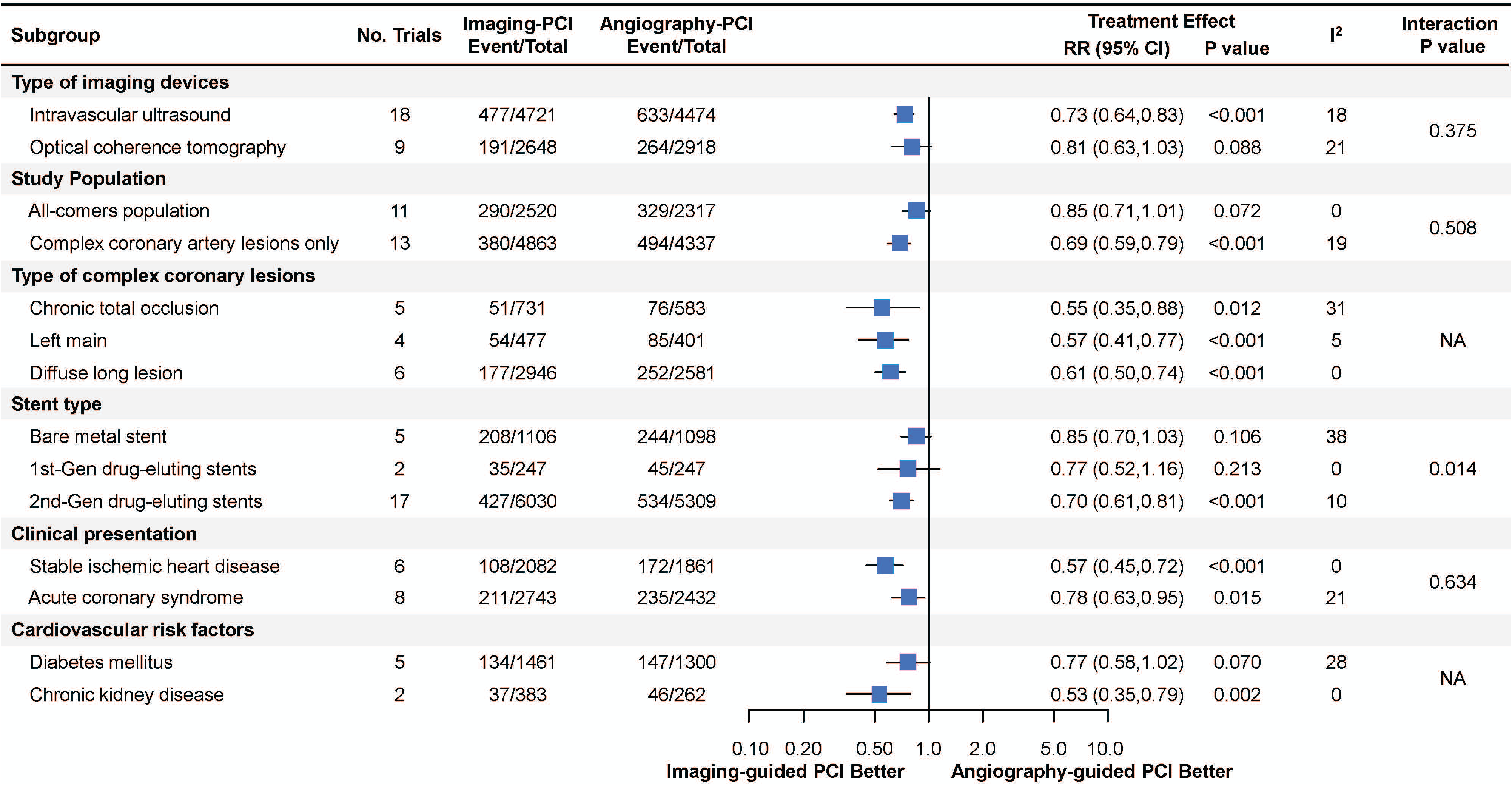
Subgroup Analysis. Forest plots of subgroup analysis comparing major adverse cardiac event, a composite of all-cause death, myocardial infarction, and target vessel revascularization between intravascular imaging-guided and angiography-guided PCI. RRs with 95% CIs are displayed for individual studies and the pooled overall effect. Abbreviations are as in Figure 2.

**Figure 5.**
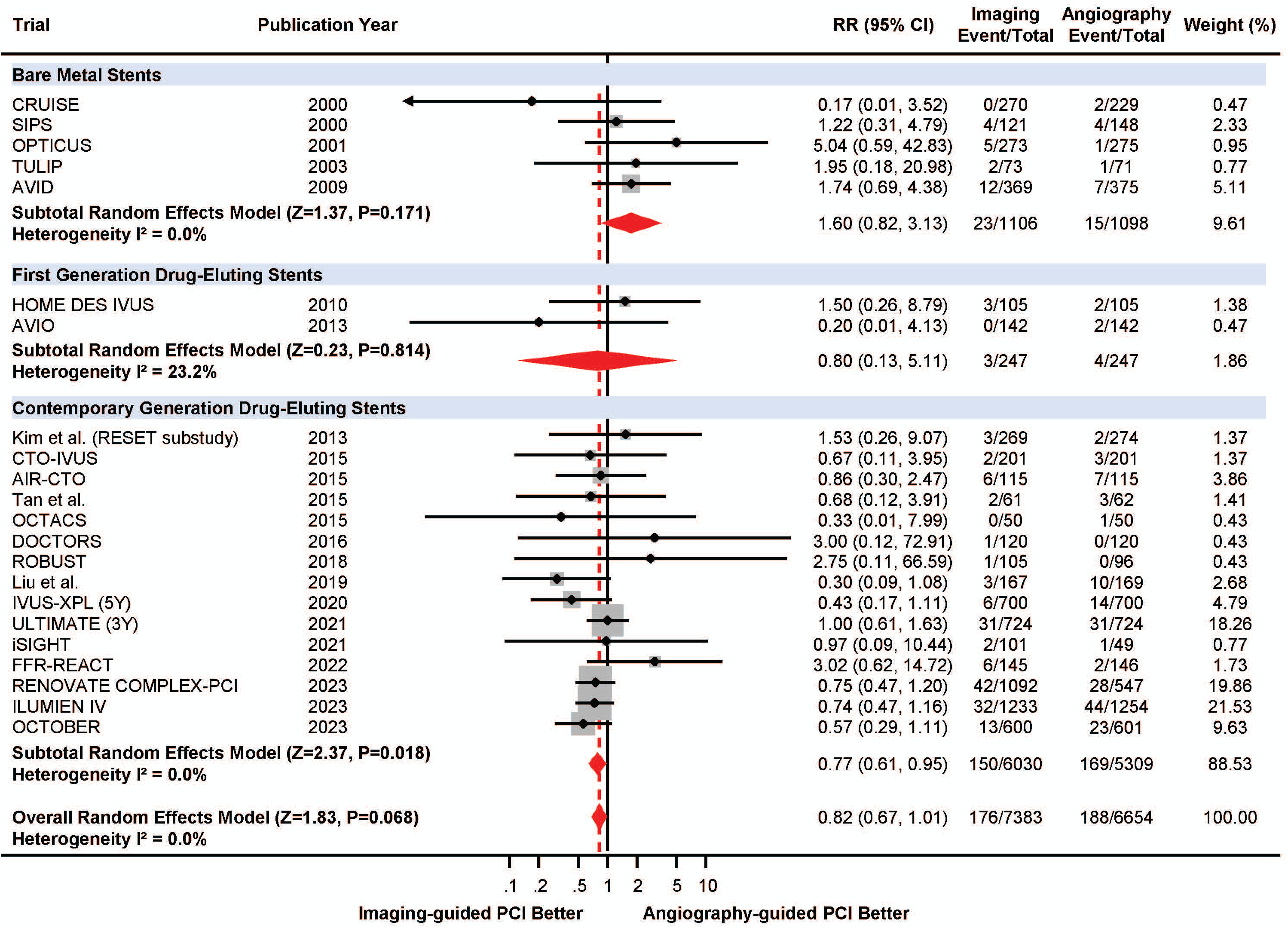
Meta-Analysis Comparing Death Between Intravascular Imaging vs. Angio-Guided Optimization Stratified by Stent Type. Forest plots comparing all-cause death between intravascular imaging-guided and angiography-guided PCI according to type of used stents. RRs with 95% CIs are displayed for individual studies and the pooled overall effect. Abbreviations are as in Figure 2.

### Patient and Public Involvement

No patient was involved in conceiving the research question, designing of the study, or interpretating the results.

## Discussion

The current updated meta-analysis compared clinical outcomes of intravascular imaging-guided PCI versus angiography-guided PCI from 24 RCTs available to date. The major findings were as follows. (1) The intravascular imaging-guided PCI significantly reduced the risk of MACE, a composite of all-cause death, MI, or TVR, compared with angiography-guided PCI. (2) The intravascular imaging-guided PCI also significantly reduced MI, TVR, and stent thrombosis, compared with angiography-guided PCI. Although overall meta-analysis showed marginal effect of intravascular imaging-guided PCI regarding all-cause death, subgroup analysis of the trials with 2^nd^ generation DES showed significantly reduced risk of all-cause death following intravascular imaging-guided PCI than angiography-guided PCI. (3) The prognostic benefit of intravascular imaging-guided PCI was consistently shown across various subgroups including complex coronary artery lesions and acute coronary syndrome.

### Current Status of Intravascular Imaging-Guided PCI

Previous RCTs have reported that intravascular imaging-guided PCI reduced the risk of MACE compared with angiography-guided PCI, largely by lowering the risk of repeat revascularization.^12–14^ ^32^ However, those RCTs have not been considered to be definitive due to limited sample size or inclusion of highly selected coronary lesion subsets. In this regard, current practice guidelines recommend the use of intravascular imaging-guided PCI as Class IIa.^1^ ^2^ Furthermore, penetration rate of intravascular imaging in daily practice has been low, although it varies across different healthcare systems or clinical indications of PCI. For example, it was approximately 7.0-9.3% in the United States^33^ ^34^ and 27.5-28.6% in South Korea.^35^

### Prognostic Impact of Intravascular Imaging-Guided PCI

Since most RCTs had only modest sample sizes to establish clinical benefit of intravascular imaging-guided PCI in terms of hard clinical events such as death, MI, or stent thrombosis, a few observational studies assessed a larger number of patients and showed that intravascular imaging-guided PCI resulted in a lower risk of death or MI than angiography-guided PCI.^33^ ^36–38^ Although previous meta-analyses presented similar results,^39^ ^40^ inclusion of both observational studies and RCTs would have caused heterogeneity and lowered the evidence level of the results. Recently, 3 large scaled RCTs have been published and showed conflicting results regarding the prognostic benefit of intravascular imaging-guided PCI.^29^ ^30^ ^41^ The RENOVATE-COMPLEX-PCI trial evaluated 1,639 patients with complex coronary artery lesions and firstly showed a superiority of imaging-guided PCI using IVUS or OCT over angiography-guided PCI in reducing the hard clinical end point including cardiac death and MI at a median follow-up of 2.1 years (7.7% vs. 12.3%, respectively; HR, 0.64; 95% CI, 0.45-0.89; P=0.008).^41^ Subsequent OCTOBER trial showed that OCT-guided PCI significantly reduced the risk of target vessel failure at 2 years than angiography-guided PCI in patients with complex bifurcation lesions (10.1% vs. 14.1%, respectively; HR, 0.70; 95% CI, 0.50-0.98; P=0.035).^29^ Conversely, ILUMIEN IV trial failed to show significant prognostic benefit of OCT-guided PCI than angiography-guided PCI in patients with diabetes mellitus or complex coronary artery lesions at 2 years (7.4% vs. 8.2%, respectively; HR, 0.90; 95% CI, 0.67-1.19; P=0.45).^30^

The current meta-analysis was performed to clarify whether intravascular imaging-guided PCI would have prognostic benefit over angiography-guided PCI, especially for hard clinical events such as all-cause death, MI, or stent thrombosis. Compared with previous meta-analyses,^39^ ^40^ the current study included only RCTs and evaluated the largest number of randomized patients. Similar to the prior results, intravascular imaging-guided PCI was associated with a lower risk of MACE compared with angiography-guided PCI among all-comers and patients with complex coronary artery lesions. The results suggested that intravascular imaging-guided PCI would prevent 1 MACE event in every 33 patients undergoing PCI. Importantly, it should be noted that intravascular imaging-guided PCI reduced not only the risk of TVR but also the risks of hard clinical events such as MI and stent thrombosis. The I² values for all clinical outcomes evaluated in the current study were less than 25%, indicating low heterogeneity among the included trials. Conversely, there was no significant difference in the risk of all-cause mortality between the 2 groups in meta-analysis of overall trials. However, it should be noted that pooled analysis of the trials with 2^nd^ generation DES showed significantly reduced risk of all-cause death following intravascular imaging-guided PCI than angiography-guided PCI. Considering that bare metal stent or 1^st^ generation DES are no longer used in contemporary PCI and that the risks of MI and stent thrombosis significantly reduced following intravascular imaging-guided PCI, it would be reasonable to accept survival benefit of intravascular imaging-guided PCI in contemporary practice. Currently ongoing trials will evaluate IVUS-guided PCI in all-comers with complex coronary artery lesions (IMPROVE [NCT04221815] and IVUS-CHIP [NCT04854070]) or unprotected left main disease (OPTIMAL [NCT04111770] and DKCRUSH-VIII [NCT03770650]), or OCT-guided PCI (OCCUPI [NCT03625908]), and these trials will provide additional information on the benefit of intravascular imaging-guided PCI, especially regarding the risk of death after PCI.

### Future Perspectives of Intravascular Imaging-Guided PCI

In the current study, the treatment effect of intravascular imaging-guided PCI was consistent regardless of clinical presentation, cardiovascular comorbidities, type of used stents, and presence of complex coronary lesions. However, it should be noted that the effect size of the intravascular imaging-guided PCI for the risk of MACE was relatively higher in patients with complex coronary artery lesions. These results were in line with the current guidelines which recommend the use of intravascular imaging-guided PCI mainly for complex coronary artery lesions.^1^ ^2^ With emerging evidences, further discussion is needed in interventional societies and guidelines committees as to whether intravascular imaging-guided PCI should be upgraded to Class I recommendation for complex coronary artery lesions. As shown in the subgroup analysis, among complex coronary artery lesions, more evidence from RCTs has been accumulated in chronic total occlusion, left main disease, and diffuse long lesion than other complex coronary artery lesions. Integration of ongoing trials and in-depth discussion are also needed to adequately define the complex coronary artery lesion that may benefit the most from intravascular imaging-guided PCI. In addition, it should not be underestimated that intravascular imaging-guided PCI also reduced the risk of MACE in all-comers. Concurrently, efforts should be made to standardize intravascular imaging optimization criteria that are practical, feasible, and prognostically well-validated. Considering low penetration rates of intravascular imaging in daily practice, more efforts need to be made to increase awareness and educate and support practicing interventional cardiologists.^42^ Furthermore, cost-effectiveness of intravascular imaging-guided PCI should be clarified to help increase its penetration rate in daily practice.

In the current meta-analysis, intravascular imaging-guided PCI significantly reduced the risk of MACE in both stable ischemic heart disease and acute coronary syndrome patients. However, there has been limited evidence from RCT regarding the role of intravascular imaging-guided PCI in acute coronary syndrome. Similarly, there have only been a few RCTs comparing the clinical benefit of OCT-guided PCI over angiography-guided PCI and recent two large scaled trials (ILUMIEN IV and OCTOBER) showed conflicting results.^29^ ^30^ To date, supporting evidence for OCT-guided PCI was mostly derived from RCTs which showed comparable clinical outcomes between IVUS- and OCT-guided PCI.^20^ ^21^ ^43^ ^44^ Similarly, subgroup analysis of the current study demonstrated no significant interaction according to the type of intravascular imaging devices, suggesting similar treatment effect between IVUS- and OCT-guided PCI. These results are in line with the recently published OCTIVUS trial which showed comparable clinical outcomes between IVUS- and OCT-guided PCI.^44^ Currently ongoing OCCUPI (NCT03625908) trial will test the potential superiority of OCT-guided PCI over angiography-guided PCI. Furthermore, since those trials are enrolling both stable ischemic heart disease and acute coronary syndrome patients, their results will further enrich the evidence supporting the value of intravascular imaging-guided PCI.

## Limitations

Some limitations should be acknowledged. First, this meta-analysis included clinically and methodologically diverse studies. Although we included only RCTs to the final analysis and got insignificant statistical inconsistency or heterogeneity, there were some differences in the study design including the enrollment criteria, type of stent used, type of intravascular imaging devices and procedural optimization criteria, follow-up protocols, and adjunctive pharmacotherapy after PCI. Second, the included trials were conducted from 2000 to 2023 and significant changes in practice patterns such as stent technology, PCI technique, and pharmacotherapy should be considered to interpret the results. However, the results from the cumulative meta-analysis imply that treatment effect of intravascular imaging-guided PCI was consistent over 20 years of period. Third, as this is a study-level meta-analysis, data of the individual patients were not included in the analysis, and therefore, we could not adjust for patient-level confounders especially the differences in target lesion complexity. Fourth, this meta-analysis basically composed of 24 RCTs inherently shares the limitations of each trial. Fifth, we did not use Google Scholar or Web of Science in the initial search strategies, and some RCTs were added manually. However, same 24 RCTs were selected even after including these databases in search strategies and the current meta-analysis included the largest number of RCTs than previously conducted meta-analyses. Sixth, as current study included only a small number of patients with ST-segment elevation MI, caution is needed to extrapolate the results to patients with ST-segment elevation MI.

## Conclusion

Compared to angiography-guided PCI, intravascular imaging-guided PCI was associated with a reduced the risk of MACE by lowering the risk of MI, TVR, and stent thrombosis. Pooled analysis of trials with 2^nd^ generation DES showed significantly lower risk of all-cause death following intravascular imaging-guided PCI than angiography-guided PCI.

## Data Availability

Original data used in the current meta-analysis including the analytic codes will be shared upon reasonable request. Any relevant inquiry should be emailed to Dr. Joo Myung Lee.
The lead author (JML) affirms that the manuscript is an honest, accurate, and transparent account of the study being reported; that no important aspects of the study have been omitted, and that any discrepancies from the study as planned have been explained.

## Acknowledgment

None.

## Contributors

DH contributed to conception and design of the work, acquisition, analysis, interpretation of data for the work, drafting of the manuscript. WK contributed to acquisition, analysis, and interpretation of data for the work. SHL contributed to the conception of data for the work, interpretation of data for the work, and critical revision of the manuscript. JML contributed to the conception and design conception and design of the work, acquisition, analysis, interpretation of data for the work, and drafting of the manuscript. All other authors contributed to interpretation of data for the work, and critical revision of the manuscript. All authors approved the final version to be published. The guarantors (DH, WK, and JML) accept full responsibility for the work and/or the conduct of the study, had access to the data, and controlled the decision to publish. The corresponding authors (SHL and JML) attest that all listed authors meet authorship criteria and that no others meeting the criteria have been omitted.

## Ethical approval

Not required.

## Data Sharing

Original data used in the current meta-analysis including the analytic codes will be shared upon reasonable request. Any relevant inquiry should be emailed to Dr. Joo Myung Lee.

The lead author (JML) affirms that the manuscript is an honest, accurate, and transparent account of the study being reported; that no important aspects of the study have been omitted, and that any discrepancies from the study as planned have been explained.

## Dissemination to participants and related patient and public communities

There are no plans to disseminate the results to the individual study participants or the relevant patient community.

## Copyright/License for Publication

The Corresponding Author (JML) has the right to grant on behalf of all authors and does grant on behalf of all authors, a worldwide licence to the Publishers and its licensees in perpetuity, in all forms, formats and media (whether known now or created in the future), to i) publish, reproduce, distribute, display and store the Contribution, ii) translate the Contribution into other languages, create adaptations, reprints, include within collections and create summaries, extracts and/or, abstracts of the Contribution, iii) create any other derivative work(s) based on the Contribution, iv) to exploit all subsidiary rights in the Contribution, v) the inclusion of electronic links from the Contribution to third party material where-ever it may be located; and, vi) licence any third party to do any or all of the above.

## Abbreviations

CI: confidence interval
DES: drug-eluting stent
IVUS: intravascular ultrasound
MACE: major adverse cardiac events
MI: myocardial infarction
OCT: optical coherence tomography
OR: odds ratio
PCI: percutaneous coronary intervention
RCT: randomized controlled trial
TVR: target vessel revascularization

