## Supplementary Appendix for "Intravascular Imaging-Guided Versus Angiography-Guided PCI: Systematic Review and Meta-analysis of 24 Randomized Controlled Trials"

David Hong, MD, Woochan Kwon, MD, Seung Hun Lee, MD, PhD, Doosup Shin, MD, Joo-Myung Lee, MD, MPH, PhD

### Table of Contents


- **Supplemental Figures and Figure Legend**
- **Supplemental Tables**

#### Supplementary Figures and Figure Legend

#### Supplemental Figure 1. Risk of Bias of Individual Trials by Cochrane Risk Assessment Tool

+ Low risk of bias
- High risk of bias
? Uncertain risk of bias

| | CRUISE | SIPS | OPTICUS | TULIP | AVID | HOME DES IVUS | AVIO | Kim et al. (RESET substudy) | CTO-IVUS | AIR-CTO | Tan et al. | OCTACS | Kim et al. | DOCTORS | ROBUST | Liu et al. | IVUS-XPL | ILUMIEN III | ULTIMATE | iSIGHT | FFR-REACT | RENOVATE-COMPLEX-PCI | ILUMIEN IV | OCTOBER |
|---|---|---|---|---|---|---|---|---|---|---|---|---|---|---|---|---|---|---|---|---|---|---|---|---|
| Random sequence generation | + | + | + | ? | + | + | ? | + | + | + | ? | + | + | + | + | + | + | + | + | + | + | + | + | + |
| Allocation Concealment | + | + | + | + | + | + | + | + | + | + | + | + | + | + | + | + | + | + | + | + | + | + | + | + |
| Blinding of participants and personnel | + | + | + | + | + | + | + | + | + | + | + | + | + | + | + | + | + | + | + | + | + | + | + | + |
| Blinding of outcome assessment | + | + | + | + | + | ? | + | + | + | + | ? | + | ? | + | + | + | + | + | + | + | + | + | + | + |
| Incomplete outcome data | + | + | + | + | + | ? | ? | + | + | + | ? | + | + | + | + | + | + | + | + | + | + | + | + | + |
| Selective reporting | + | + | + | + | + | + | + | + | + | + | + | + | + | + | + | + | + | + | + | + | + | + | + | + |
| Other bias | + | + | + | + | + | + | + | + | + | + | + | + | + | + | + | + | + | + | + | + | + | + | + | + |

Risk of bias assessment according to the Cochrane Collaboration's tool (RoB 2.0) for randomized controlled trials.
Abbreviations: **AIR-CTO**, Angiographic and clinical comparisons of intravascular ultrasound- versus angiography-guided drug-eluting stent implantation for patients with chronic total occlusion lesions; **AVID**, Angiography Versus Intravascular ultrasound-Directed stent placement; **AVIO**, Angiography Vs. IVUS Optimization; **CRUISE**, Can Routine Ultrasound Influence Stent Expansion; **CTO-IVUS**, Chronic Total Occlusion InterVention with drUg-eluting Stents guided by IVUS; **DOCTORS**, Does Optical Coherence Tomography Optimize Results of Stenting; **FFR-REACT**, Fractional flow reserve guided percutaneous coronary intervention optimization directed by high-definition intravascular ultrasound versus standard of care; **HOME DES IVUS**, Long-Term Health Outcome and Mortality Evaluation After Invasive Coronary Treatment Using Drug Eluting Stents with or without the IVUS Guidance; **iSIGHT**, Optical Coherence Tomography Versus Intravascular Ultrasound and Angiography to Guide Percutaneous Coronary Interventions; **IVUS-XPL**, Impact of Intravascular Ultrasound Guidance on Outcomes of Xience Prime Stents in Long Lesions; **OCTACS**, Optical Coherence Tomography Guided Percutaneous Coronary

Intervention With Nobori Stent Implantation in Patients With Non ST Segment Elevation Myocardial Infarction; **OCTOBER**, European Trial on Optical Coherence Tomography Optimized Bifurcation Event Reduction; **OPTICUS**, OPTimization with ICUS to reduce stent restenosis; **RENOVATE COMPLEX-PCI**, Randomized Controlled Trial of Intravascular Imaging Guidance versus Angiography-Guidance on Clinical Outcomes After Complex Percutaneous Coronary Intervention; **RESET**, Real Safety and Efficacy of a 3-Month Dual Antiplatelet Therapy Following Zotarolimus-Eluting Stents Implantation; **ROBUST**, Comparison of Biolimus A9 and Everolimus Drug-Eluting Stents in Patients With ST Segment Elevation Myocardial Infarction; **SIPS**, Strategy for Intracoronary Ultrasound-Guided PTCA and Stenting; **TULIP**, Thrombocyte activity evaluation and effects of Ultrasound guidance in Long Intracoronary stent Placement; and **ULTIMATE**, Intravascular Ultrasound Guided Drug Eluting Stents Implantation in “All-Comers” Coronary Lesions.

**Supplemental Figure 2. Cumulative Meta-Analysis**

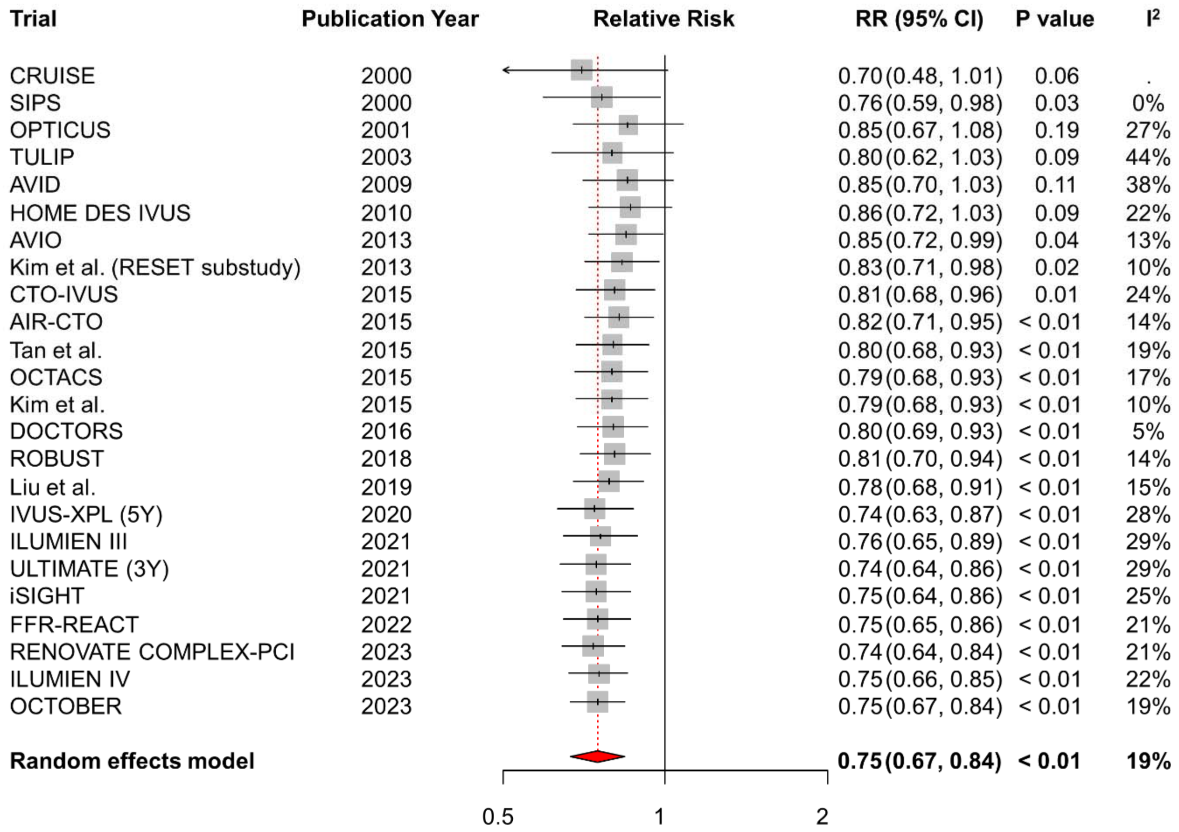


Cumulative meta-analysis for the association between major adverse cardiac event and intravascular imaging-guided percutaneous coronary intervention across overall study period from 2000 to 2023.

Abbreviations: CI, confidence interval; RR, relative risk; and other abbreviations are as in **Supplemental Figure 1**.

**Supplemental Figure 3. Influence of Individual Study**

| Trial | Publication Year | RR (95% CI) | P value | $I^2$ |
|---|---|---|---|---|
| CRUISE | 2000 | 0.75 (0.66, 0.85) | < 0.01 | 22% |
| SIPS | 2000 | 0.74 (0.66, 0.84) | < 0.01 | 22% |
| OPTICUS | 2001 | 0.73 (0.65, 0.82) | < 0.01 | 10% |
| TULIP | 2003 | 0.76 (0.68, 0.85) | < 0.01 | 17% |
| AVID | 2009 | 0.73 (0.65, 0.82) | < 0.01 | 12% |
| HOME DES IVUS | 2010 | 0.75 (0.66, 0.84) | < 0.01 | 22% |
| AVIO | 2013 | 0.75 (0.66, 0.85) | < 0.01 | 22% |
| Kim et al. (RESET substudy) | 2013 | 0.75 (0.67, 0.85) | < 0.01 | 21% |
| CTO-IVUS | 2015 | 0.76 (0.67, 0.85) | < 0.01 | 16% |
| AIR-CTO | 2015 | 0.74 (0.66, 0.84) | < 0.01 | 22% |
| Tan et al. | 2015 | 0.76 (0.67, 0.85) | < 0.01 | 18% |
| OCTACS | 2015 | 0.75 (0.67, 0.84) | < 0.01 | 20% |
| Kim et al. | 2015 | 0.75 (0.67, 0.84) | < 0.01 | 22% |
| DOCTORS | 2016 | 0.75 (0.67, 0.84) | < 0.01 | 21% |
| ROBUST | 2018 | 0.75 (0.66, 0.84) | < 0.01 | 14% |
| Liu et al. | 2019 | 0.76 (0.67, 0.85) | < 0.01 | 20% |
| IVUS-XPL (5Y) | 2020 | 0.77 (0.69, 0.86) | < 0.01 | 9% |
| ILUMIEN III | 2021 | 0.74 (0.66, 0.83) | < 0.01 | 17% |
| ULTIMATE (3Y) | 2021 | 0.76 (0.67, 0.86) | < 0.01 | 18% |
| iSIGHT | 2021 | 0.75 (0.67, 0.84) | < 0.01 | 22% |
| FFR-REACT | 2022 | 0.75 (0.67, 0.84) | < 0.01 | 22% |
| RENOVATE COMPLEX-PCI | 2023 | 0.76 (0.67, 0.86) | < 0.01 | 19% |
| ILUMIEN IV | 2023 | 0.74 (0.65, 0.83) | < 0.01 | 17% |
| OCTOBER | 2023 | 0.75 (0.66, 0.85) | < 0.01 | 22% |
| **Random effects model** | | **0.75 (0.67, 0.84)** | **< 0.01** | **19%** |

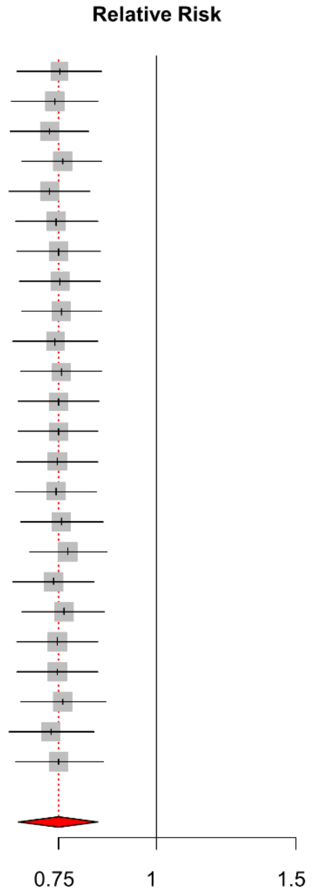


Sensitivity analyses for the influence of individual study on the pooled RRs. The circles and the horizontal lines indicate the RRs (by random effects model) and the 95% CI for each trial excluded.

Abbreviations are as in **Supplemental Figure 1 and 2**.

**Supplemental Figure 4. Assessment of Small Study Bias**

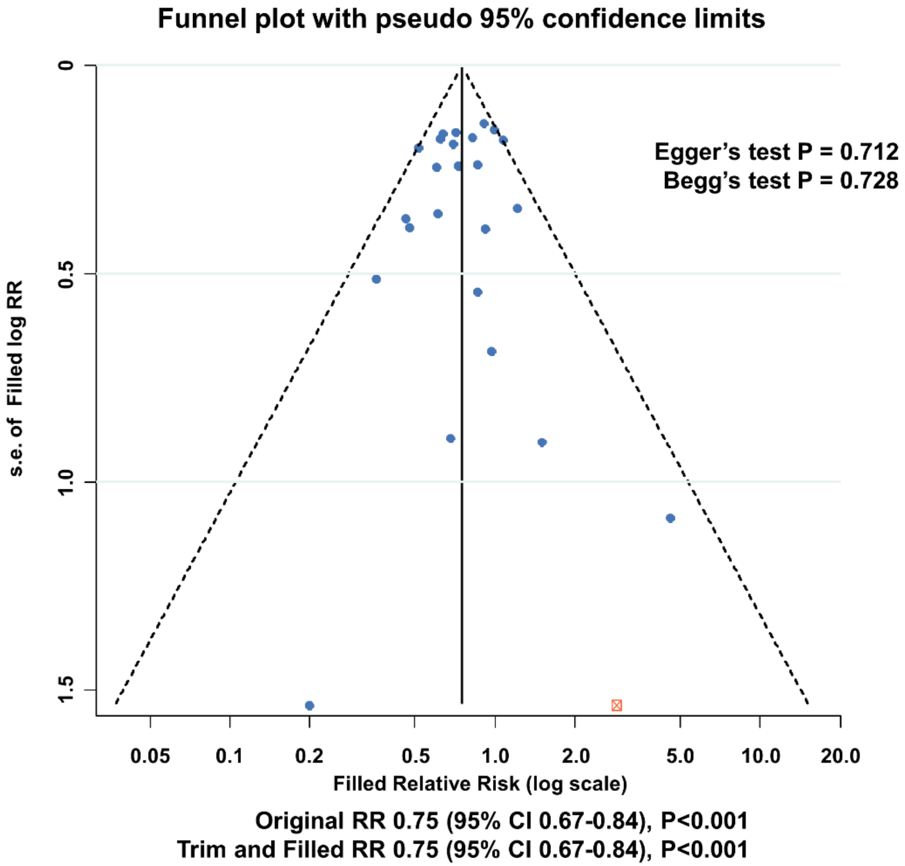


The results of Egger's and Begg's tests are presented. When appropriate, the estimated relative risk, trim-and-fill method, and the original relative risk are shown.

Abbreviations are as in **Supplemental Figure 2**.

**Supplemental Figure 5. Meta-Analysis Comparing Major Adverse Cardiac Event Between Intravascular Imaging vs. Angio-Guided Optimization Stratified by Clinical Presentations**

**A. Stable Ischemic Heart Disease**

| Trial | Publication Year | RR (95% CI) | Imaging Events/Total | Angiography Events/Total | Weight (%) |
|---|---|---|---|---|---|
| CTO-IVUS | 2015 | 0.36 (0.13, 0.97) | 5/201 | 14/201 | 5.4 |
| IVUS-XPL (5Y) | 2020 | 0.47 (0.26, 0.85) | 15/358 | 32/356 | 15.4 |
| ULTIMATE (3Y) | 2021 | 0.41 (0.13, 1.26) | 4/155 | 10/157 | 4.2 |
| RENOVATE COMPLEX-PCI | 2023 | 0.48 (0.28, 0.81) | 25/532 | 27/275 | 19.8 |
| ILUMIEN IV | 2023 | 0.75 (0.48, 1.17) | 31/506 | 45/551 | 28.0 |
| OCTOBER | 2023 | 0.62 (0.40, 0.97) | 28/330 | 44/321 | 27.1 |
| **Overall Random Effects Model (P<0.001; I² = 0.0%)** | | **0.57 (0.45, 0.72)** | **108/2082** | **172/1861** | **100.0** |

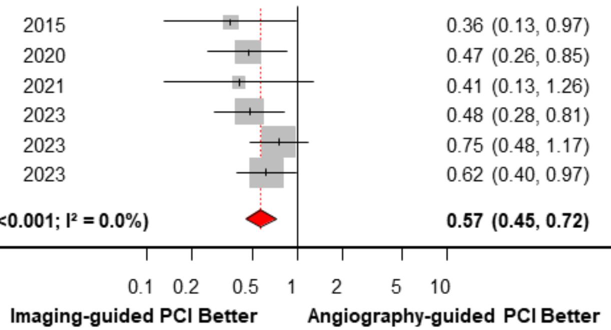


**B. Acute Coronary Syndrome**

| Trial | Publication Year | RR (95% CI) | Imaging Events/Total | Angiography Events/Total | Weight (%) |
|---|---|---|---|---|---|
| OCTACS | 2015 | 0.20 (0.01, 4.06) | 0/50 | 2/50 | 0.5 |
| DOCTORS | 2016 | 1.50 (0.26, 8.82) | 3/120 | 2/120 | 1.3 |
| ROBUST | 2018 | 4.57 (0.54, 38.43) | 5/105 | 1/96 | 0.9 |
| IVUS-XPL (5Y) | 2020 | 0.56 (0.33, 0.93) | 21/342 | 38/344 | 13.5 |
| ULTIMATE (3Y) | 2021 | 0.65 (0.45, 0.94) | 43/569 | 66/567 | 23.2 |
| RENOVATE COMPLEX-PCI | 2023 | 0.75 (0.50, 1.13) | 51/560 | 33/272 | 19.2 |
| ILUMIEN IV | 2023 | 1.02 (0.71, 1.46) | 57/727 | 54/703 | 24.1 |
| OCTOBER | 2023 | 0.82 (0.53, 1.28) | 31/270 | 39/280 | 17.3 |
| **Overall Random Effects Model (P=0.015; I² = 21.0%)** | | **0.78 (0.63, 0.95)** | **211/2743** | **235/2432** | **100.0** |

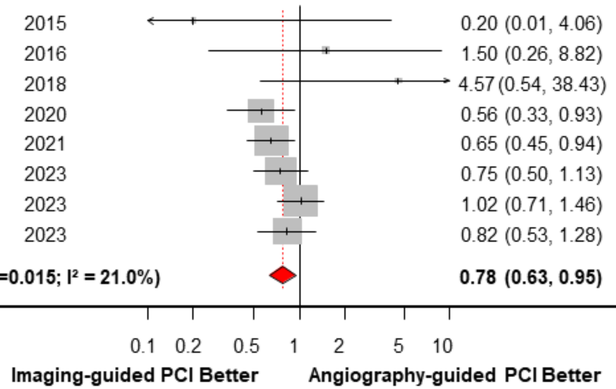


Forest plots comparing major adverse cardiac event, a composite of all-cause death, myocardial infarction, and target vessel revascularization between intravascular imaging-guided and angiography-guided PCI stratified by clinical presentation. (A) stable ischemic heart disease and (B) acute coronary syndrome.

Abbreviations are as in **Supplemental Figure 1 and 2**.

### Supplementary Tables

#### Supplemental Table 1. Search Strategy

| Pubmed | | | EMBASE | | | Cochrane Library* | | |
|---|---|---|---|---|---|---|---|---|
| **#15** | #14 Filter: Randomized Clinical Trial | **281** | **#15** | #14 AND randomized | **185** | **#15** | #14 AND randomized | **343** |
| **#14** | #8 AND #13 | **2194** | **#14** | #8 AND #13 | **858** | **#14** | #8 AND #13 | **416** |
| **#13** | #6 AND #12 | **57749** | **#13** | #6 AND #12 | **38493** | **#13** | #6 AND #12 | **10427** |
| **#12** | #9 OR #10 OR #11 | **4285655** | **#12** | #9 OR #10 OR #11 | **3730409** | **#12** | #9 OR #10 OR #11 | **788382** |
| **#11** | Outcomes | **3113310** | **#11** | Outcomes | **2133597** | **#11** | Outcomes | **714897** |
| **#10** | Impact | **1516353** | **#10** | Impact | **1922592** | **#10** | Impact | **145141** |
| **#9** | Outcome | **3113310** | **#9** | Outcome | **3220160** | **#9** | Outcome | **714871** |
| **#8** | #3 AND #7 | **19593** | **#8** | #3 AND #7 | **30897** | **#8** | #3 AND #7 | **1923** |
| **#7** | angiography | **345646** | **#7** | angiography | **446520** | **#7** | angiography | **17825** |
| **#6** | #4 or #5 | **108334** | **#6** | #4 or #5 | **122716** | **#6** | #4 or #5 | **15342** |
| **#5** | Percutaneous coronary intervention | **79047** | **#5** | Percutaneous coronary intervention | **122620** | **#5** | Percutaneous coronary intervention | **12428** |
| **#4** | Stent implantation | **39160** | **#4** | Stent implantation | **72** | **#4** | Stent implantation | **4630** |
| **#3** | #1 or #2 | **75770** | **#3** | #1 or #2 | **102396** | **#3** | #1 or #2 | **6261** |
| **#2** | Optical coherence tomography [Title/Abstract] | **62218** | **#2** | Optical coherence tomography [Title/Abstract] | **87293** | **#2** | Optical coherence tomography | **4884** |
| **#1** | Intravascular ultrasound [Title/Abstract] | **14944** | **#1** | Intravascular ultrasound [Title/Abstract] | **18092** | **#1** | Intravascular ultrasound | **1538** |

*The number of results refer to the number of search results exclusively from Trial section.

**Supplemental Table 2. Reasons of Excluded Studies After Full Article Review**

| Source (Year) | Study Name | Acronym | Reason of Exclusion |
|---|---|---|---|
| 2011 | Impact of intravascular ultrasound imaging on early and late clinical outcomes following percutaneous coronary intervention with drug-eluting stents | | Irrelevant design (Cohort study with prospective enrolled registry) |
| 2019 | Intravascular ULTrasound Guided Versus Conventional Angiography Guided Strategy to Deploy Zotarolimus and Everolimus Eluting Third Generation Stents in the Long Coronary Artery Lesions: ULTRA-ZET Trial | ULTRA-ZET | No published outcome data due to early termination |
| 2020 | Intravascular Ultrasound Versus Angiography Guided Drug-coated Balloon Treatment for STEMI Patients – Prospective, Multicenter, Randomized Controlled Trial | | No available outcome data (Not published yet) |
| 2003 | Continued improvement of clinical outcome and cost effectiveness following intravascular ultrasound guided PCI: insights from a prospective, randomised study | | Inadequate primary endpoint – study on cost-effectiveness |
| 2007 | Comparison of angiographically guided direct stenting technique with direct stenting and optimal balloon angioplasty guided with intravascular ultrasound. The multicenter, randomized trial results. | DIPOL trial substudy | Irrelevant design – comparison of direct stenting and balloon angioplasty besides IVUS vs. angiography |
| 1998 | Impact of intravascular ultrasound guidance in stent deployment on 6-month restenosis rate: a multicenter, randomized study comparing two strategies--with and without intravascular ultrasound guidance. RESIST Study Group. REStenosis after Ivus guided STenting. | RESIST study | Irrelevant study design |
| 2013 | Usefulness of intravascular ultrasound to predict outcomes in short-length lesions treated with drug-eluting stents. | RESET trial substudy | Duplicate study population with RESET trial |
| 2021 | [Long-term outcomes of intravascular ultrasound-guided drug-eluting stent implantation in patients with chronic kidney disease: ULTIMATE CKD subgroup analysis]. | ULTIMATE trial substudy | Duplicate study population with ULTIMATE trial, article in Chinese |
| 2016 | Does Optical Coherence Tomography Optimise | DOCTORS | Duplicate results |
| 2015 | Effect of Intravascular Ultrasound-Guided vs Angiography-Guided Everolimus-Eluting Stent Implantation: The IVUS-XPL Randomized Clinical Trial. | IVUS-XPL | Duplicate study population with IVUS-XPL trial (original publication) |

| | | | |
|---|---|---|---|
| 2016 | ILUMIEN III: OPTIMIZE PCI OPtical Coherence Tomography (OCT) Compared to Intravascular Ultrasound (IVUS) and Angiography to Guide Coronary Stent Implantation: a Multicenter RandomIZEd Trial in PCI | ILUMIEN III | Duplicate study population with ILUMIEN III trial (original publication) |
| 2019 | Impact of intravascular ultrasound-guided drug-eluting stent implantation on patients with chronic kidney disease: results from ULTIMATE trial | ULTIMATE trial substudy | Duplicate study population with ULTIMATE trial |
| 2018 | Intravascular Ultrasound Versus Angiography-Guided Drug-Eluting Stent Implantation: the ULTIMATE Trial | ULTIMATE | Duplicate study population with ULTIMATE III trial (original publication) |
| 2016 | Optical coherence tomography compared with intravascular ultrasound and with angiography to guide coronary stent implantation (ILUMIEN III: OPTIMIZE PCI): a randomised controlled trial. | ILUMIEN III | Duplicate result |

**Supplemental Table 3. Definition of Major Adverse Cardiac Events in Included Trials**

| Trial | Publication Year | Definition |
|---|---|---|
| CRUISE | 2000 | Composite of death, myocardial infarction, or target-vessel revascularization |
| SIPS | 2000 | Composite of death, myocardial infarction, or repeat revascularization |
| OPTICUS | 2001 | Composite of death, myocardial infarction, or repeat revascularization |
| TULIP | 2003 | Composite of death, myocardial infarction, or target-lesion revascularization |
| AVID | 2009 | Composite of death, myocardial infarction, target-lesion revascularization, stent thrombosis, or coronary artery bypass graft |
| HOME DES IVUS | 2010 | Composite of death, myocardial infarction, or target-lesion revascularization |
| AVIO | 2013 | Composite of cardiac death, myocardial infarction, or target-lesion revascularization |
| Kim et al. (RESET substudy) | 2013 | Composite of cardiovascular death, myocardial infarction, or target-vessel revascularization or stent thrombosis |
| CTO-IVUS | 2015 | Composite of cardiac death, myocardial infarction, or target-vessel revascularization |
| AIR-CTO | 2015 | Not reported |
| Tan et al. | 2015 | Composite of death, myocardial infarction, or target-lesion revascularization |
| OCTACS | 2015 | Composite of cardiac death, myocardial infarction, repeat revascularization, or stent thrombosis |
| Kim et al. | 2015 | Composite of cardiac death, nonfatal myocardial infarction, target-lesion revascularization, or stent thrombosis |
| DOCTORS | 2016 | Composite of death, myocardial infarction, or target-vessel revascularization or stent thrombosis |
| ROBUST | 2018 | Composite of death, myocardial infarction, or target-lesion revascularization |
| Liu et al. | 2019 | Composite of cardiac death, myocardial infarction, or target-vessel revascularization |
| IVUS-XPL | 2020 | Composite of cardiac death, target-lesion related myocardial infarction, or ischemia-driven target-lesion revascularization |
| ILUMIEN III | 2021 | Composite of death, myocardial infarction, or repeat revascularization or stent thrombosis |
| ULTIMATE | 2021 | Composite of cardiac death, target-vessel myocardial infarction, or clinically driven target-vessel revascularization |
| iSIGHT | 2021 | Composite of cardiac death, myocardial infarction, or target-lesion revascularization |
| FFR-REACT | 2022 | Composite of cardiac death, spontaneous target-vessel myocardial infarction, or |

| | | |
|---|---|---|
| | | clinically driven target-vessel revascularization |
| RENOVATE-COMPLEX-PCI | 2023 | Composite of cardiac death, target-vessel-related myocardial infarction, or clinically driven target-vessel revascularization |
| ILUMIEN IV | 2023 | Composite of cardiac death, target-vessel myocardial infarction, or ischemia-driven target-vessel revascularization |
| OCTOBER | 2023 | Composite of cardiac death, target-lesion myocardial infarction, or ischemia-driven target-lesion revascularization |

**Supplemental Table 4.** The Cochrane Collaboration's tool for assessing risk of bias of 4 randomised clinical trials in meta-analysis

| Study | Domain | Support for judgment & review authors' judgment |
|---|---|---|
| **Fitzerald, P. J. et al. ("CRUISE study")** | Random Sequence Generation | Low risk of bias. The trial was a substudy of another randomized clinical trial ("STARS trial") that randomly allocated patients in equal proportion to either asprin, ticlopidine or warfarin group although the specific manner of random sequence generation was not reported. |
| | Allocation concealment | Low risk of bias. Although maintaining allocation concealment of participants and medical personnel is inherently impossible due to the characteristics of the procedure, clinical outcomes were adjudicated by independent clinical events committee, which was blinded to the treatment received. |
| | Blinding of participants and personnel | Low risk of bias. The blinding of participants and medical personnel is inherently impossible due to the characteristics of the procedure. However, all primary and secondary endpoints were objective findings, and in the angiography-guided PCI group, post-PCI IVUS measurement was done in a blinded fashion, preventing response from the operator according to the result. |
| | Blinding of outcome assessment | Low risk of bias. All end points were reviewed by the independent clinical events committee which was blinded to the treatment received. All ultrasound images were reviewed and evaluated by an independent core laboratory. |
| | Incomplete outcome data | Low risk of bias. 26 patients (5.0% of originally enrolled subjects) were lost to the follow-up. Other patients in both groups were completely followed to the end of the study and had complete adjudication for clinical follow-up. |
| | Selective reporting | Low risk of bias. All of the study's pre-specified (primary and secondary) outcomes that are of interest in the review have been reported in the pre-specified manner. |
| | Other sources of bias | Low risk of bias. The study appears to be free of other sources of bias. |

| | | |
|---|---|---|
| **Frey A. W. et al. ("SIPS study")** | Random Sequence Generation | Low risk of bias. Patients were randomized on a day-to-day block schedule at the specific point of time of the day. The specific manner of random sequence generation was not reported. |
| | Allocation concealment | Low risk of bias. Maintaining allocation concealment of participants and medical personnel is inherently impossible due to the characteristics of the procedure. The review author judged that lack of allocation concealment is not likely to influence to the results of the current study. |
| | Blinding of participants and personnel | Low risk of bias. The blinding of participants and medical personnel is inherently impossible due to the characteristics of the procedure. However, since all primary and secondary endpoints were objective findings, the review author judged that the outcome is not likely to be influenced by lack of blinding. |
| | Blinding of outcome assessment | Low risk of bias. Angiograms were analyzed by an independent core laboratory, and the frames for analysis were chosen by a physician not involved in the performance of the procedure, though the characteristic appearance of IVUS precluded complete blinding. |
| | Incomplete outcome data | Low risk of bias. Though angiographic follow-up was completed in 77% of angiography-guided group and 79% of IVUS-guided group, clinical follow-up was complete in 100% of the population in which the outcome data of interest was gathered. |
| | Selective reporting | Low risk of bias. All of the study's pre-specified outcomes that are of interest in the review have been reported in the pre-specified manner. |
| | Other sources of bias | Low risk of bias. The study appears to be free of other sources of bias. |
| **Aversano, T. et al. ("OPTICUS study")** | Random Sequence Generation | Low risk of bias. Randomization was performed by fax from an independent center (a central office at Munich University) before the start of the procedure with subrandomization performed for de novo or restenotic lesions, although the |

| | | |
|---|---|---|
| | | specific fashion of randomization was not reported. |
| | Allocation concealment | Low risk of bias. Maintaining allocation concealment of participants and medical personnel is inherently impossible due to the characteristics of the procedure. The review author judged that lack of allocation concealment is not likely to influence to the results of the current study. |
| | Blinding of participants and personnel | Low risk of bias. The blinding of participants and medical personnel is inherently impossible due to the characteristics of the procedure. However, since all primary and secondary endpoints were objective findings, the review author judged that the outcome is not likely to be influenced by lack of blinding. |
| | Blinding of outcome assessment | Low risk of bias. All angiograms were analyzed by an independent core laboratory (Frankfurt University core laboratory) blinded for treatment allocation. |
| | Incomplete outcome data | Low risk of bias. Analysis were done in intention-to-treat fashion, and the clinical follow-up loss occurred in only 2% at 6-month and 5% of the total population at 12-month point. |
| | Selective reporting | Low risk of bias. All of the study's pre-specified outcomes that are of interest in the review have been reported in the pre-specified manner. |
| | Other sources of bias | Low risk of bias. The study appears to be free of other sources of bias. |
| **Oemrawsingh, P. V. et al. ("TULIP study")** | Random Sequence Generation | Unclear risk of bias. Randomization was performed just before the procedure, although the specific fashion of randomization was not reported. |
| | Allocation concealment | Low risk of bias. Maintaining allocation concealment of participants and medical personnel is inherently impossible due to the characteristics of the procedure. The review author judged that lack of allocation concealment is not likely to influence to the results of the current study. |
| | Blinding of participants and | Low risk of bias. The blinding of participants and medical personnel is inherently |

| | | |
|---|---|---|
| | personnel | impossible due to the characteristics of the procedure. However, since all primary and secondary endpoints were objective findings, the review author judged that the outcome is not likely to be influenced by lack of blinding. |
| | Blinding of outcome assessment | Low risk of bias. All angiograms were analyzed by an independent core laboratory (HeartCore, Leiden, the Netherlands) blinded as to IVUS assignment |
| | Incomplete outcome data | Low risk of bias. Follow-up loss was kept to minimal (4%), which was lower than expected rate of follow-up loss (10-15%). |
| | Selective reporting | Low risk of bias. All of the study's pre-specified outcomes that are of interest in the review have been reported in the pre-specified manner. |
| | Other sources of bias | Low risk of bias. The study appears to be free of other sources of bias. |
| **Russo, R.J. et al ("AVID study")** | Random Sequence Generation | Low risk of bias. Balanced, blocked randomization was used to allocate equal numbers of subjects at each site to Angiography- or IVUS-directed stent placement. Computer-generated treatment assignments were placed in serially numbered, sealed, opaque envelopes by the coordinating center. |
| | Allocation concealment | Low risk of bias. Maintaining allocation concealment of participants and medical personnel is inherently impossible due to the characteristics of the procedure. The review author judged that lack of allocation concealment is not likely to influence to the results of the current study. |
| | Blinding of participants and personnel | Low risk of bias. The blinding of participants and medical personnel is inherently impossible due to the characteristics of the procedure. However, since all primary and secondary endpoints were objective findings, the review author judged that the outcome is not likely to be influenced by lack of blinding. |
| | Blinding of outcome assessment | Low risk of bias. Treating physicians who performed stent placement and IVUS imaging and cardiac catheterization laboratory staff were not blinded due to the characteristics. However, patients and staff members of the angiography and |

| | | |
|---|---|---|
| | | IVUS core laboratories (Washington Hospital Center Angiographic Core Laboratory) who performed image measurements, nurses who conducted telephone follow-up, and physicians at the coordinating center who adjudicated outcome events were blinded to the treatment allocation assignment. |
| | Incomplete outcome data | Low risk of bias. Only 7% of patients were lost to follow-up, which was lower than the anticipated rate of follow-up loss for sample size calculation. |
| | Selective reporting | Low risk of bias. All of the study's pre-specified outcomes that are of interest in the review have been reported in the pre-specified manner. |
| | Other sources of bias | Low risk of bias. The study appears to be free of other sources of bias. |
| **Jakabcin J. et al ("HOME DES IVUS study")** | Random Sequence Generation | Low risk of bias. Randomization employing sealed envelope was performed in a 1:1 ratio and patients were assigned to the two treatment groups. |
| | Allocation concealment | Low risk of bias. Maintaining allocation concealment of participants and medical personnel is inherently impossible due to the characteristics of the procedure. The review author judged that lack of allocation concealment is not likely to influence to the results of the current study. |
| | Blinding of participants and personnel | Low risk of bias. The blinding of participants and medical personnel is inherently impossible due to the characteristics of the procedure. However, since all primary and secondary endpoints were objective findings, the review author judged that the outcome is not likely to be influenced by lack of blinding. |
| | Blinding of outcome assessment | Unclear risk of bias. The exact nature of blinding method is not reported, though it was implied ultrasound images were sent for off-line analysis in a separate laboratory. |
| | Incomplete outcome data | Unclear risk of bias. The rate of follow-up loss is not reported, though the analysis is done on the premise that follow-up data was available in every |

| | | |
|---|---|---|
| | | enrolled subjects. |
| | Selective reporting | Low risk of bias. All of the study's pre-specified outcomes that are of interest in the review have been reported in the pre-specified manner. |
| | Other sources of bias | Low risk of bias. The study appears to be free of other sources of bias. |
| **Chieffo A. et al ("AVIO study")** | Random Sequence Generation | Unclear risk of bias. Patients were randomized at the time of the angiogram, through sealed opaque envelopes. |
| | Allocation concealment | Low risk of bias. Maintaining allocation concealment of participants and medical personnel is inherently impossible due to the characteristics of the procedure. The review author judged that lack of allocation concealment is not likely to influence to the results of the current study. |
| | Blinding of participants and personnel | Low risk of bias. The blinding of participants and medical personnel is inherently impossible due to the characteristics of the procedure. However, since all primary and secondary endpoints were objective findings, the review author judged that the outcome is not likely to be influenced by lack of blinding. |
| | Blinding of outcome assessment | Low risk of bias. Data handling and adjudication were performed in an independent center (Mediolanum Cardio Research, Milan, Italy) blinded to a group allocation |
| | Incomplete outcome data | Unclear risk of bias. The rate of follow-up loss is not reported, though the analysis is done on the premise that follow-up data was available in every enrolled subjects. |
| | Selective reporting | Low risk of bias. All of the study's pre-specified outcomes that are of interest in the review have been reported in the pre-specified manner. |
| | Other sources of bias | Low risk of bias. The study appears to be free of other sources of bias. |

| | | |
|---|---|---|
| **Kim, JS et al. ("RESET substudy")** | Random Sequence Generation | Low risk of bias. Balanced, blocked randomization was conducted via a web-based randomization system. |
| | Allocation concealment | Low risk of bias. Maintaining allocation concealment of participants and medical personnel is inherently impossible due to the characteristics of the procedure. The review author judged that lack of allocation concealment is not likely to influence to the results of the current study. |
| | Blinding of participants and personnel | Low risk of bias. The blinding of participants and medical personnel is inherently impossible due to the characteristics of the procedure. However, since all primary and secondary endpoints were objective findings, the review author judged that the outcome is not likely to be influenced by lack of blinding. |
| | Blinding of outcome assessment | Low risk of bias. Angiographic and ultrasound data analysis was performed by individuals who were blinded to treatment assignment in an independent core laboratory at Cardiovascular Research Center, Seoul, Korea. |
| | Incomplete outcome data | Low risk of bias. Though the crossover to the other assigned treatment group occurred in 54 patients, per-protocol analysis was also done and produced similar results. Clinical follow-up was completed in all patients for pre-specified study duration. |
| | Selective reporting | Low risk of bias. All of the study's pre-specified outcomes that are of interest in the review have been reported in the pre-specified manner. |
| | Other sources of bias | Low risk of bias. The study appears to be free of other sources of bias. |
| **Kim, BK et al ("CTO-IVUS study")** | Random Sequence Generation | Low risk of bias. Study participants were randomly assigned and stratified by participating center using an interactive web-based response system. Allocations were generated using a permuted-block randomization with varying block sizes. |
| | Allocation concealment | Low risk of bias. Maintaining allocation concealment of participants and medical personnel is inherently impossible due to the characteristics of the procedure. |

| | | |
|---|---|---|
| | | The review author judged that lack of allocation concealment is not likely to influence to the results of the current study. |
| | Blinding of participants and personnel | Low risk of bias. The blinding of participants and medical personnel is inherently impossible due to the characteristics of the procedure. However, since all primary and secondary endpoints were objective findings, the review author judged that the outcome is not likely to be influenced by lack of blinding. |
| | Blinding of outcome assessment | Low risk of bias. All clinical events were independently monitored and assessed by a clinical event committee blinded to randomization. Angiographic and IVUS data analysis were done by analysts blinded to the details of the patient and procedural information. |
| | Incomplete outcome data | Low risk of bias. Only 1 patient was lost to follow-up and other 401 patients completed 12-month follow-up. |
| | Selective reporting | Low risk of bias. All of the study's pre-specified outcomes that are of interest in the review have been reported in the pre-specified manner. |
| | Other sources of bias | Low risk of bias. The study appears to be free of other sources of bias. |
| **Tian, NL et al. ("AIR-CTO study")** | Random Sequence Generation | Low risk of bias. Though more detailed method of randomization was not reported, randomization was done by randomization program of the central computer. |
| | Allocation concealment | Low risk of bias. Maintaining allocation concealment of participants and medical personnel is inherently impossible due to the characteristics of the procedure. The review author judged that lack of allocation concealment is not likely to influence to the results of the current study. |
| | Blinding of participants and personnel | Low risk of bias. The blinding of participants and medical personnel is inherently impossible due to the characteristics of the procedure. However, since all primary and secondary endpoints were objective findings, the review author |

| | | |
|---|---|---|
| | | judged that the outcome is not likely to be influenced by lack of blinding. |
| | Blinding of outcome assessment | Low risk of bias. Though the on-site assessment of post-procedural IVUS images was performed by experienced technicians, the analyses of angiograms and IVUS images were performed by core lab technicians (Nanjing Heart Centre, Nanjing, China) who were blinded to the study design |
| | Incomplete outcome data | Low risk of bias. Though angiographic and IVUS follow-up were done in 50-70% of the subjects, clinical follow-up was done on all but only 4 patients, and the review author judged that the high rate of follow-up is adequate for inclusion in the current study. |
| | Selective reporting | Low risk of bias. All of the study's pre-specified outcomes that are of interest in the review have been reported in the pre-specified manner. |
| | Other sources of bias | Low risk of bias. The study appears to be free of other sources of bias. |
| **Tan, Q. et al** | Random Sequence Generation | Unclear risk of bias. Patients were randomized into two groups, though the exact method of randomization was not reported. |
| | Allocation concealment | Low risk of bias. Maintaining allocation concealment of participants and medical personnel is inherently impossible due to the characteristics of the procedure. The review author judged that lack of allocation concealment is not likely to influence to the results of the current study. |
| | Blinding of participants and personnel | Low risk of bias. The blinding of participants and medical personnel is inherently impossible due to the characteristics of the procedure. However, since all primary and secondary endpoints were objective findings, the review author judged that the outcome is not likely to be influenced by lack of blinding. |
| | Blinding of outcome assessment | Unclear risk of bias. Blinding method is not detailed, though it is implied that angiogram and IVUS data analysis were done in an independent core laboratory. |

| | | |
|---|---|---|
| | Incomplete outcome data | Unclear risk of bias. The rate of follow-up loss is not reported, though the analysis is done on the premise that follow-up data was available in every enrolled subjects. |
| | Selective reporting | Low risk of bias. All of the study's pre-specified outcomes that are of interest in the review have been reported in the pre-specified manner. |
| | Other sources of bias | Low risk of bias. The study appears to be free of other sources of bias. |
| **Kim et al** | Random Sequence Generation | Low risk of bias. Patients were randomly assigned in a 1:1 ratio by an interactive web-based response system of participating centers. Randomization was stratified according to the presence of diabetes mellitus, acute coronary syndrome, and the estimated length and diameter of the prospective drug-eluting stent implant. |
| | Allocation concealment | Low risk of bias. Maintaining allocation concealment of participants and medical personnel is inherently impossible due to the characteristics of the procedure. The review author judged that lack of allocation concealment is not likely to influence to the results of the current study. |
| | Blinding of participants and personnel | Low risk of bias. The blinding of participants and medical personnel is inherently impossible due to the characteristics of the procedure. However, since all primary and secondary endpoints were objective findings, the review author judged that the outcome is not likely to be influenced by lack of blinding. |
| | Blinding of outcome assessment | Unclear risk of bias. Blinding method is not detailed, though it is implied that angiogram and OCT data analysis were done in an independent core laboratory. |
| | Incomplete outcome data | Low risk of bias. All patients completed 12-month clinical follow-up. |
| | Selective reporting | Low risk of bias. All of the study's pre-specified outcomes that are of interest in the review have been reported in the pre-specified manner. |

| | | |
|---|---|---|
| | Other sources of bias | Low risk of bias. The study appears to be free of other sources of bias. |
| **Antonsen, L. et al ("OCTACS trial")** | Random Sequence Generation | Low risk of bias. Random assignments to the treatment groups were distributed in sealed envelopes. |
| | Allocation concealment | Low risk of bias. Maintaining allocation concealment of participants and medical personnel is inherently impossible due to the characteristics of the procedure. The review author judged that lack of allocation concealment is not likely to influence to the results of the current study. |
| | Blinding of participants and personnel | Low risk of bias. Though it was not possible to blind the operator, investigator, or patient for the allocated implantation technique, the operator was blinded to the postprocedure OCT images, because the operator screen-side was turned off, and the entire pullback remained uncommented on. |
| | Blinding of outcome assessment | Low risk of bias. The operator was blinded to the postprocedure OCT images, and the OCT analyst of the core laboratory who analyzed the OCT images was blinded to the implantation technique, when analyzing 6-month pullbacks. |
| | Incomplete outcome data | Low risk of bias. Though invasive follow-up was lost in 24.0% of the patients, clinical follow-up was complete in all patients. |
| | Selective reporting | Low risk of bias. All of the study's pre-specified outcomes that are of interest in the review have been reported in the pre-specified manner. |
| | Other sources of bias | Low risk of bias. The study appears to be free of other sources of bias. |
| **Meneveau, N. et al ("DOCTORS study")** | Random Sequence Generation | Low risk of bias. Patients were randomized to two groups after initial coronary angiography stratified by center. Randomization was performed using consecutive sealed opaque envelopes. |
| | Allocation concealment | Low risk of bias. Maintaining allocation concealment of participants and medical personnel is inherently impossible due to the characteristics of the procedure. The review author judged that lack of allocation concealment is not likely to |

| | | |
|---|---|---|
| | | influence to the results of the current study. |
| | Blinding of participants and personnel | Low risk of bias. The blinding of participants and medical personnel is inherently impossible due to the characteristics of the procedure. However, since all primary and secondary endpoints were objective findings, the review author judged that the outcome is not likely to be influenced by lack of blinding. |
| | Blinding of outcome assessment | Low risk of bias. All OCT and angiography images were analyzed in a centralized core laboratory (University Hospital of Besancon) by 2 independent operators blinded to the angiographic findings, procedural strategy, and final FFR value |
| | Incomplete outcome data | Low risk of bias. Only one patient was lost to follow-up, whose survival state was identified by municipal death registries. Sample size calculation was done expecting at least five follow-up loss. |
| | Selective reporting | Low risk of bias. All of the study's pre-specified outcomes that are of interest in the review have been reported in the pre-specified manner. Formal data monitoring was overseen by the Clinical Research Management Department of the coordinating center by sending independent monitors to each site regularly to monitor files and check data entry. |
| | Other sources of bias | Low risk of bias. The study appears to be free of other sources of bias. |
| **Ali, Z.A. et al ("ILUMIEN III study")** | Random Sequence Generation | Low risk of bias. Randomization was done with use of an interactive web-based system in block sizes of three, stratified by site. |
| | Allocation concealment | Low risk of bias. Maintaining allocation concealment of participants and medical personnel is inherently impossible due to the characteristics of the procedure. The review author judged that the outcome is not likely to be influenced by lack of blinding. |

| | | |
|---|---|---|
| | Blinding of participants and personnel | Low risk of bias. The blinding of participants and medical personnel is inherently impossible due to the characteristics of the procedure. However, post-PCI stent area, assessed by OCT, were masked to the operators in IVUS and angiography group, partially concealing the allocation. However, since all primary and secondary endpoints were objective findings, the review author judged that the outcome is not likely to be influenced by lack of blinding. |
| | Blinding of outcome assessment | Low risk of bias. The independent core laboratory that analysed the primary outcome and the independent clinical events committee that adjudicated clinical and safety events were also masked to treatment assignment |
| | Incomplete outcome data | Low risk of bias. Only 9 patients were lost to follow-up, and per-protocol analysis was also done that produced the similar results. |
| | Selective reporting | Low risk of bias. All of the study's pre-specified outcomes that are of interest in the review have been reported in the pre-specified manner. |
| | Other sources of bias | Low risk of bias. The study appears to be free of other sources of bias. |
| **Kala, P. et al ("ROBUST trial substudy")** | Random Sequence Generation | Low risk of bias. Patients were randomly assigned using a sealed envelope after diagnostic coronary angiography. |
| | Allocation concealment | Low risk of bias. Maintaining allocation concealment of participants and medical personnel is inherently impossible due to the characteristics of the procedure. The review author judged that lack of allocation concealment is not likely to influence to the results of the current study. |
| | Blinding of participants and personnel | Low risk of bias. The blinding of participants and medical personnel is inherently impossible due to the characteristics of the procedure. However, since all primary and secondary endpoints were objective findings, the review author judged that the outcome is not likely to be influenced by lack of blinding. |
| | Blinding of outcome assessment | Low risk of bias. The images were recorded in the OCT system console and |

| | | |
|---|---|---|
| | | analyzed on-line in the cathlab and off-line in independent CoreLab. |
| | Incomplete outcome data | Low risk of bias. Almost 90% of the patients were available for follow-up OCT and clinical follow-up loss was not reported. |
| | Selective reporting | Low risk of bias. All of the study's pre-specified outcomes that are of interest in the review have been reported in the pre-specified manner. |
| | Other sources of bias | Low risk of bias. The study appears to be free of other sources of bias. |
| **Liu, XM et al.** | Random Sequence Generation | Low risk of bias. Opaque envelopes written with different IDs indicating the related groups were used to randomly divide the enrolled patients. |
| | Allocation concealment | Low risk of bias. Maintaining allocation concealment of participants and medical personnel is inherently impossible due to the characteristics of the procedure. The review author judged that lack of allocation concealment is not likely to influence to the results of the current study. |
| | Blinding of participants and personnel | Low risk of bias. The blinding of participants and medical personnel is inherently impossible due to the characteristics of the procedure. However, since all primary and secondary endpoints were objective findings, the review author judged that the outcome is not likely to be influenced by lack of blinding. |
| | Blinding of outcome assessment | Low risk of bias. An independent cardiologist blinded to the study was in charge of assessing all events. |
| | Incomplete outcome data | Low risk of bias. Only 9 patients were lost to follow-up and its effect on accuracy was deemed minimal. |
| | Selective reporting | Low risk of bias. All of the study's pre-specified outcomes that are of interest in the review have been reported in the pre-specified manner. |
| | Other sources of bias | Low risk of bias. The study appears to be free of other sources of bias. |

| | | |
|---|---|---|
| **Hong, SJ et al. ("IVUS-XPL trial")** | Random Sequence Generation | Low risk of bias. Study participants were randomly assigned in a 1:1 ratio to two groups immediately after coronary angiography but before percutaneous coronary intervention, though the specific method of randomization was not reported. |
| | Allocation concealment | Low risk of bias. Maintaining allocation concealment of participants and medical personnel is inherently impossible due to the characteristics of the procedure. The review author judged that lack of allocation concealment is not likely to influence to the results of the current study. |
| | Blinding of participants and personnel | Low risk of bias. The blinding of participants and medical personnel is inherently impossible due to the characteristics of the procedure. However, since all primary and secondary endpoints were objective findings, the review author judged that the outcome is not likely to be influenced by lack of blinding. |
| | Blinding of outcome assessment | Low risk of bias. A blinded independent clinical events committee adjudicated all nonprocedural components of the primary endpoint on the basis of the original source documents. |
| | Incomplete outcome data | Low risk of bias. Only 6% of patients were lost to 1-year follow-up. 15% were lost to 5-year follow-up but the statistical power was deemed adequate. |
| | Selective reporting | Low risk of bias. All of the study's pre-specified outcomes that are of interest in the review have been reported in the pre-specified manner. |
| | Other sources of bias | Low risk of bias. The study appears to be free of other sources of bias. |
| **Gao, XF et al. ("ULTIMATE study")** | Random Sequence Generation | Low risk of bias. Patients were randomized in a 1:1 ratio to receive either IVUS or angiography guidance by random envelope method before PCI. A matched block method stratified by clinicians was used to generate random sequence of envelop allocations. |
| | Allocation concealment | Low risk of bias. Maintaining allocation concealment of participants and medical |

| | | |
|---|---|---|
| | | personnel is inherently impossible due to the characteristics of the procedure. The review author judged that lack of allocation concealment is not likely to influence to the results of the current study. |
| | Blinding of participants and personnel | Low risk of bias. The blinding of participants and medical personnel is inherently impossible due to the characteristics of the procedure. However, since all primary and secondary endpoints were objective findings, the review author judged that the outcome is not likely to be influenced by lack of blinding. |
| | Blinding of outcome assessment | Low risk of bias. An independent events committee who was blinded to study design and randomization results assessed all clinical events. |
| | Incomplete outcome data | Low risk of bias. Only four patients were lost in 12-month follow-up, and additional 21 patients lost in 3-year point, which only consists of 1.7% of total population. |
| | Selective reporting | Low risk of bias. All of the study's pre-specified outcomes that are of interest in the review have been reported in the pre-specified manner. |
| | Other sources of bias | Low risk of bias. The study appears to be free of other sources of bias. |
| **Chamie, D. et al. ("iSIGHT trial")** | Random Sequence Generation | Low risk of bias. The randomization sequence was electronically generated in block sizes of 9 and secured in opaque sealed envelopes, opened after guidewire positioning. |
| | Allocation concealment | Low risk of bias. Maintaining allocation concealment of participants and medical personnel is inherently impossible due to the characteristics of the procedure. However, to reduce bias, patients in the angiography arm were given blinded IVUS and OCT at the end of the procedure, and patients in the IVUS and OCT arms were given the opposite imaging modality. |
| | Blinding of participants and personnel | Low risk of bias. The blinding of participants and medical personnel is inherently impossible due to the characteristics of the procedure. However, since all |

| | | |
|---|---|---|
| | | primary and secondary endpoints were objective findings, the review author judged that the outcome is not likely to be influenced by lack of blinding. |
| | Blinding of outcome assessment | Low risk of bias. At the end of each procedure, all images were deidentified and transferred to the core laboratories, where all analysts were blinded to the randomization groups. |
| | Incomplete outcome data | Low risk of bias. Only one patient was lost to follow-up and the outcome data was available in almost all subjects. |
| | Selective reporting | Low risk of bias. All of the study's pre-specified outcomes that are of interest in the review have been reported in the pre-specified manner. |
| | Other sources of bias | Low risk of bias. The study appears to be free of other sources of bias. |
| **Neleman, T. et al ("FFR-REACT trial")** | Random Sequence Generation | Low risk of bias. Randomization (block size varying from 4-6) was performed online in a 1:1 fashion by a web-based application. |
| | Allocation concealment | Low risk of bias. Maintaining allocation concealment of participants and medical personnel is inherently impossible due to the characteristics of the procedure. The review author judged that lack of allocation concealment is not likely to influence to the results of the current study. |
| | Blinding of participants and personnel | Low risk of bias. The blinding of participants and medical personnel is inherently impossible due to the characteristics of the procedure. However, since all primary and secondary endpoints were objective findings, the review author judged that the outcome is not likely to be influenced by lack of blinding. |
| | Blinding of outcome assessment | Low risk of bias. All FFR tracings and IVUS pull backs were assessed off-line in a blinded fashion by the Erasmus University Medical Center academic core laboratory. Patients, physicians involved in patient care, study personnel performing follow-up calls and visits, and the independent clinical event committee were blinded to post-PCI FFR values and group allocation. Per |

| | | |
|---|---|---|
| | | protocol, operators were uninvolved in the study follow-up and analysis. |
| | Incomplete outcome data | Low risk of bias. Only two patients were lost to follow-up and the outcome data was available in almost all subjects. |
| | Selective reporting | Low risk of bias. All of the study's pre-specified outcomes that are of interest in the review have been reported in the pre-specified manner. |
| | Other sources of bias | Low risk of bias. The study appears to be free of other sources of bias. |
| **Lee, JM et al. ("RENOVATE-COMPLEX-PCI study")** | Random Sequence Generation | Low risk of bias. Randomization was performed by a web-based randomization program developed by an independent organization and was stratified by clinical presentation and participating centers. |
| | Allocation concealment | Low risk of bias. Maintaining allocation concealment of participants and medical personnel is inherently impossible due to the characteristics of the procedure. The review author judged that lack of allocation concealment is not likely to influence to the results of the current study. |
| | Blinding of participants and personnel | Low risk of bias. The blinding of participants and medical personnel is inherently impossible due to the characteristics of the procedure. However, since all primary and secondary endpoints were objective findings, the review author judged that the outcome is not likely to be influenced by lack of blinding. |
| | Blinding of outcome assessment | Low risk of bias. All angiograms and intravascular imaging data were analyzed in the independent core laboratories. |
| | Incomplete outcome data | Low risk of bias. Only one patient was lost to follow-up and the outcome data was available in almost all subjects. |
| | Selective reporting | Low risk of bias. All of the study's pre-specified outcomes that are of interest in the review have been reported in the pre-specified manner. |

| | | |
|---|---|---|
| | Other sources of bias | Low risk of bias. The study appears to be free of other sources of bias. |
| **Holm, N.R. et al (“OCTOBER trial”)** | Random Sequence Generation | Low risk of bias. Randomization was performed with the use of a concealed, external, web-based randomization service. The randomization sequence was performed in permuted blocks in random sizes of 4, 6, and 8 and was stratified according to the presence of a left main bifurcation lesion and types of the planned stenting techniques. |
| | Allocation concealment | Low risk of bias. Maintaining allocation concealment of participants and medical personnel is inherently impossible due to the characteristics of the procedure. The review author judged that lack of allocation concealment is not likely to influence to the results of the current study. |
| | Blinding of participants and personnel | Low risk of bias. The blinding of participants and medical personnel is inherently impossible due to the characteristics of the procedure. However, since all primary and secondary endpoints were objective findings, the review author judged that the outcome is not likely to be influenced by lack of blinding. |
| | Blinding of outcome assessment | Low risk of bias. angiograms and OCT scans from the index procedure and staged procedures were submitted to the trial core laboratory (Aarhus University, Denmark). Members of an independent clinical-end-point committee who were unaware of the trial group assignments adjudicated all definite and possible clinical events. |
| | Incomplete outcome data | Low risk of bias. Only 2% of the study population were lost to the follow up, and all but five cases’ angiogram were missing vital parts for analysis. |
| | Selective reporting | Low risk of bias. All of the study’s pre-specified outcomes that are of interest in the review have been reported in the pre-specified manner. |
| | Other sources of bias | Low risk of bias. The study appears to be free of other sources of bias. |
| **Ali, Z.A. et al** | Random Sequence Generation | Low risk of bias. Randomization was performed in variable block sizes and was |

| | | |
|---|---|---|
| **("ILUMIEN IV study")** | | stratified according to medication-treated diabetes mellitus, presentation with a myocardial infarction, and trial site. Exact method of randomization was not recorded. |
| | Allocation concealment | Low risk of bias. Maintaining allocation concealment of participants and medical personnel is inherently impossible due to the characteristics of the procedure. The review author judged that lack of allocation concealment is not likely to influence to the results of the current study. |
| | Blinding of participants and personnel | Low risk of bias. Subjects were blinded to their treatment assignment and the study site personnel were trained not to disclose the treatment assignment to the subject. In addition to standard procedural sedation, headphones were worn by the patient during the procedure to reduce the possibility of unblinding. In addition, any records the patient may have access to did not refer to details of intravascular imaging or use other revealing language, to maintain the blinding. |
| | Blinding of outcome assessment | Low risk of bias. Blinded site personnel, not present at the index procedure conducted the clinical follow-up. |
| | Incomplete outcome data | Low risk of bias. Only 6% of patients were lost to follow-up, which was approximately the estimated number of those lost to follow-up in sample size calculation. For the principal analyses, missing data were not replaced; however, as a sensitivity analysis, multiple imputation was used to account for missing data. |
| | Selective reporting | Low risk of bias. All of the study's pre-specified outcomes that are of interest in the review have been reported in the pre-specified manner. |
| | Other sources of bias | Low risk of bias. The study appears to be free of other sources of bias. |

Abbreviations. PCI, percutaneous coronary intervention; IVUS, intravascular ultrasound; OCT, optical coherence tomography; FFR, fractional flow reserve.

**Supplemental Table 5. Baseline Characteristics of Patients in Included Trials**

| Trial | Age, y | Male, % | Hypertension, % | Diabetes Mellitus, % | Dyslipidemia, % | Acute Coronary Syndrome, % |
|---|---|---|---|---|---|---|
| CRUISE | 60/61 | 69/72 | 52/59 | 23/18 | 39/33 | NR |
| SIPS | 61/61 | 82/76 | 64/56 | 16/16 | 88/87 | NR |
| OPTICUS | 60/62 | 77/78 | 48/52 | 17/17 | 61/67 | NR |
| TULIP | 61/63 | 71/72 | 27/30 | 16/21 | 61/62 | NR |
| AVID | 62/63 | 73/68 | 46/45 | 15/17 | 40/44 | NR |
| HOME DES IVUS | 60/60 | 73/71 | 67/71 | 42/45 | 63/66 | 72/60 |
| AVIO | 64/64 | 82/77 | 70/67 | 24/27 | 70/77 | NR |
| Kim et al. (RESET substudy) | 63/64 | 66/55 | 61/66 | 32/30 | 61/62 | 47/49 |
| CTO-IVUS | 61/61 | 81/81 | 63/64 | 35/34 | NR | 0/0 |
| AIR-CTO | 67/66 | 89/80 | 75/70 | 30/27 | 22/28 | 29/24 |
| Tan et al. | 77/76 | 62/69 | 41/47 | 34/30 | NR | 71/66 |
| Kim et al. | 59/62 | 39/37 | 27/25 | 16/16 | 33/37 | 19/20 |
| OCTACS | 62/67 | 72/68 | 56/56 | 16/10 | 44/38 | 100/100 |
| DOCTORS | 61/61 | 79/76 | 56/42 | 22/16 | 49/47 | 100/100 |
| ROBUST | 57/59 | 83/87 | 50/52 | 17/26 | NR | 100/100 |
| Liu et al. | 65/65 | 64/64 | 70/72 | 34/31 | 38/38 | 86/87 |
| IVUS-XPL | 64/64 | 69/69 | 65/63 | 36/37 | 67/65 | 49/49 |
| ILUMIEN III | 66/67 | 71/73 | 78/75 | 35/28 | 74/77 | 35/36 |
| ULTIMATE | 65/66 | 74/73 | 71/72 | 30/31 | 54/55 | 79/78 |
| iSIGHT | 60/59 | 66/78 | 85/80 | 37/45 | 65/57 | 60/57 |
| FFR-REACT | 66/67 | 85/77 | 70/73 | 26/18 | 69/61 | 44/51 |
| RENOVATE COMPLEX-PCI | 65/66 | 80/79 | 63/59 | 36/41 | 51/51 | 51/50 |
| ILUMIEN IV | 66/66 | 79/76 | 71/74 | 42/42 | 66/69 | 59/56 |
| OCTOBER | 66/66 | 89/89 | 70/75 | 17/16 | 76/79 | 45/47 |

Data are presented as intravascular imaging-guided PCI/angiography-guided PCI.
Abbreviations: NR, not presented; PCI, percutaneous coronary intervention.
